# Evolutionarily conserved pathways of caregiving breakdown underlie contemporary child maltreatment

**DOI:** 10.64898/2026.07.03.26357058

**Authors:** Yuko Shiraishi, Eri Miyazawa, Kumi O Kuroda

**Author notes:** These authors were equally contributed to this study (Yuko Shiraishi, Eri Miyazawa). Corresponding author: Kumi O. Kuroda School of Life Science and Technology, Institute of Science Tokyo 4259 Nagatsuta-cho, Midori-ku, Yokohama, Kanagawa 226-8503, Japan.

## Abstract

Child maltreatment is a leading cause of preventable harm, yet how diverse risk factors accumulate to precipitate caregiving failure remains unclear. In non-human mammals, fatal abandonment or aggression toward offspring occurs under specific ecological conditions, suggesting evolutionarily conserved pathways for caregiving breakdown. To test whether similar structures apply to humans, we conducted a case–control study enrolling 39 caregivers imprisoned for (near-) lethal child maltreatment and 351 control caregivers in Japan. Across 70 examined factors spanning caregivers’ childhood, socioeconomic, neurobiological, and proximate environmental profiles, severe maltreatment was associated with 4.6- and 5.9-fold higher exposure to cross-species factors in men and women, respectively. Developmental pathway modeling identified significant standardized total effects of low educational attainment, non-kin caregiving, isolated parenting, behavioral addiction, and maternal absence before age 15. This proof-of-concept study presents an integrated framework bridging evolutionary biology with contemporary human caregiving and suggests targets for actionable intervention.

**Research highlights:**

- Severe child maltreatment in a modern society mirrors evolutionarily conserved, “cross-species” risk structures for caregiving breakdown.
- Early-life adversities precede isolated parenting and non-kin caregiving, the most prominent proximal predictors of maltreatment.
- The absence of a same-gender genetic parent during childhood exerts a greater effect size on later maltreatment than opposite-gender parent absence.
- Suggesting targeted parenting support for multi-risk families and educational support for children in adversity.

## Introduction

Child maltreatment remains a profound global challenge, compromising children’s well-being throughout life and imposing a long-term societal burden through the intergenerational cycle of violence ^1–3^. While cumulative risks across caregiver, child, household, and community levels have been associated with child maltreatment ^4–6^, caregivers’ early-life adversities are found as the most frequent predictors ^6–8^. Yet, the complex nature of caregivers’ life-course modeling ^9^ hindered our ability to clarify how diverse risk factors accumulate to precipitate caregiving failure.

An evolutionary perspective may help disentangle the etiology of severe human maltreatment. Parental care is mandatory for all mammalian infants, and its neural and behavioral systems are highly conserved across species ^10,11^. Nevertheless, lethal offspring desertion (neglect) or aggression toward offspring (physical abuse) occurs under specific ecological conditions, suggesting the existence of evolutionarily conserved pathways leading to caregiving breakdown.

We previously proposed that in non-human mammals, caregiving failure can be categorized into “non-adaptive” and “adaptive” contexts ^12^. Non-adaptive failure typically arises from insufficient social learning due to premature reproduction or early-life adversity, or from direct neurobiological impairment (Table 1, i-ii) ^10,11^. In contrast, “adaptive” abandonment or strategic allocation of limited resources emerges when parents face ecological constraints or excessive offspring demands (Environmental, iii-2, 3). This represents an evolutionary mechanism to prioritize long-term reproductive success over unviable current investment. In addition, “adaptive” infanticide is observed when animals sacrifice genetically unrelated infants to prioritize investment in biological offspring (iii-1) ^13^. While each of these adaptive and non-adaptive causes has been investigated for humans independently ^14,15^, empirical research that comprehensively covers these cross-species risks remains scarce. This is at least partly because in-depth information about the caregiver is limited in public records, such as child death reviews or court documents ^16,17^.

**Table 1.**
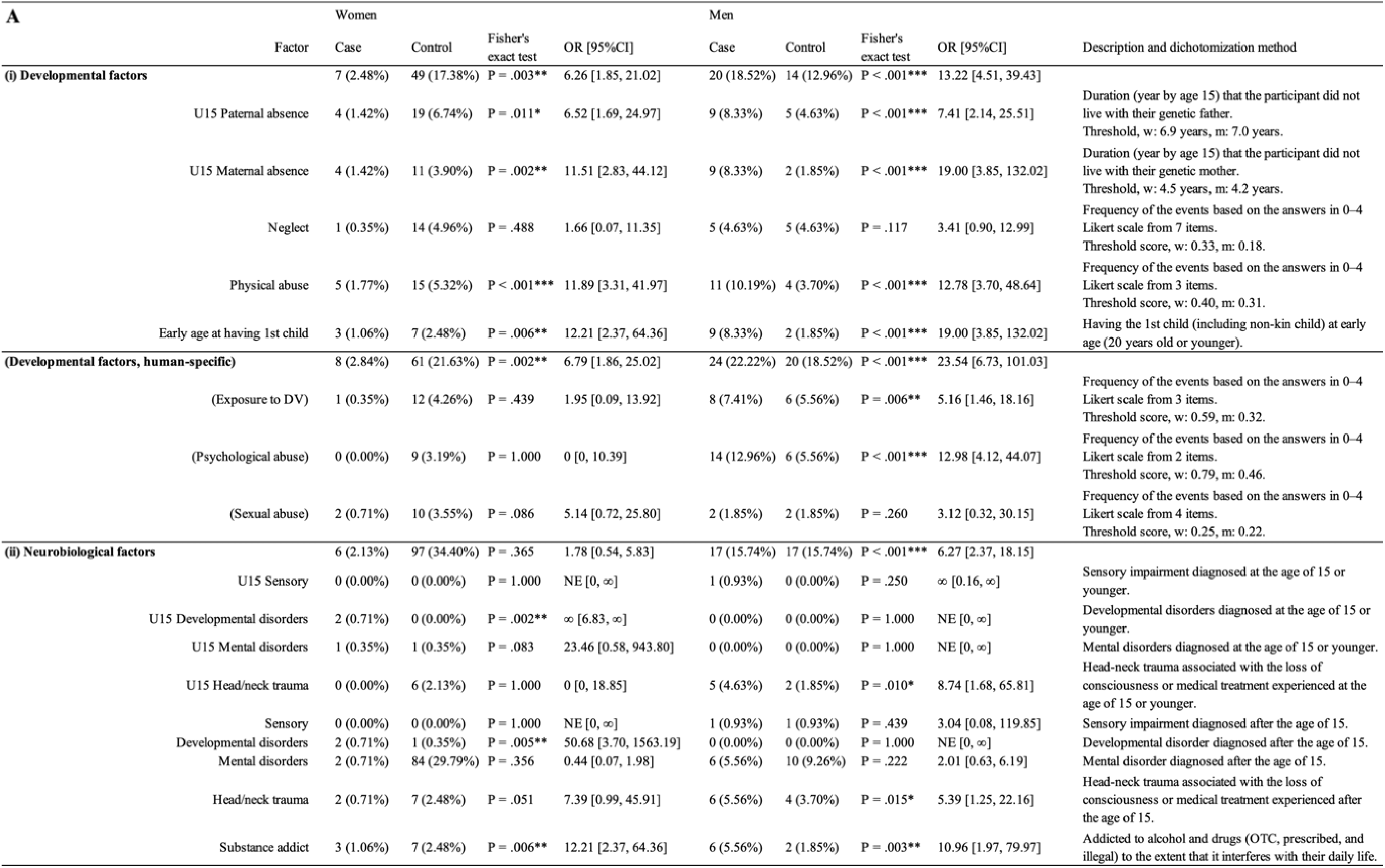
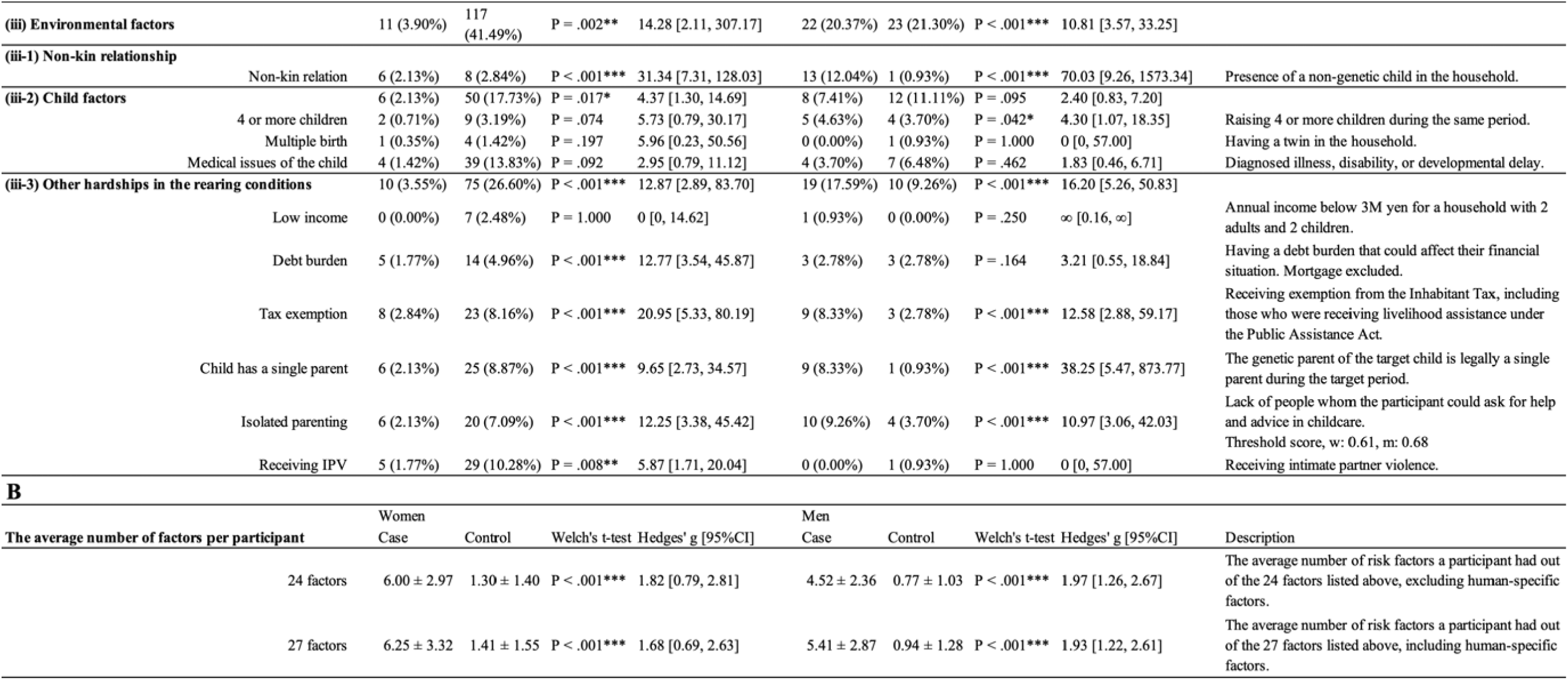
Group comparisons of cross-species factors for caregiving failure. **A.** human counterparts of the established risk factors for mammalian parenting across five domains ^12^: (i) developmental, (ii) neurobiological, and (iii) environmental, further subdivided into (iii-1) non-kin relation, (iii-2) child factors, (iii-3) socio-ecological hardships during parenthood. The number and percentage of participants who met each factor are shown. Odds ratios and 95% confidence intervals were estimated using the Control group as the reference. NE: not estimable. **B.** The average number of risk factors per participant. Mean ± SD for the number of risk factors (24 factors plus 3 types of maltreatment that have not been well studied in animals) applicable to participants in each group are shown. Note: primiparity (the mother being primiparous for the target child) is a known risk factor for maternal desertion of offspring in mammals; however, it was not included in this study for the following reasons: first, it does not apply to men, and second, Control participants with multiple children were instructed to report on their latest child. Asterisks denote statistical significance based on Fisher’s exact test (A) or Welch’s t-test (B), comparing the Case and Control groups (* P < .05, ** P < .01, *** P < .001).

To address this gap and obtain detailed information directly from maltreatment perpetrators, here we recruited 390 Japanese adults with caregiving experience, including 39 caregivers incarcerated for severe maltreatment and a control group of caregivers. From 402 firsthand data items supplemented by public records, we identified 24 factors that exhibit conceptual correspondence with established inhibitors of parenting in non-human mammals, hereafter designated as “cross-species” risk factors. This study integrates these factors with human-specific variables to test how evolutionarily conserved mechanisms of caregiving breakdown apply to a contemporary human society.

## Results

### The Case group

Using databases of major Japanese newspapers, we identified the names and court sentences of 302 adults convicted of child maltreatment between 2006 and 2018. Invitation letters were sent via postal mail to 139 convicts estimated to be serving their sentences at that time. Of the 86 individuals who were successfully reached, 48 (55.8%) replied with a written interest in participating. Ultimately, 39 participants (81.3% of those consenting, 27 men and 12 women) completed the entire survey and were included in the Case group.

The demographic details of these Case participants are summarized in Table 2. Most maltreatment cases involved physical abuse (82.1%), followed by neglect (12.8%) (Fig. S1A). The mean age of the victimized children is 3.0 ± 3.0 (0-14) years old (Fig. S1B). Regarding biological relatedness, 91.7 % (11 of 12) of the female convicts and 51.9% (14 of 27) of the male convicts were the (reportedly) genetic parents of the victimized child. The remaining convicts were non-kin adults to the victimized child, including 6 stepfathers and 6 boyfriends of the genetic mothers. These non-kin adults were grouped together in this study because Daly and colleagues reported that non-kinship is the major risk factor of severe child abuse regardless of legal relationship ^18^(see ^19^ however). Comparisons between our Case group and Child Maltreatment Fatality Reviews (CMFR) in Japan between 2010 and 2022 (hereafter the CMFR cases) ^17^ suggest that our Case group reasonably represents severe child maltreatment cases in Japan (Fig. S1), except for early-infancy neglect by young genetic mothers, who are more likely to receive probation.

**Table 2.** Characteristics of the participants and the target child. Age at the time of responding to the questionnaire and years of post-compulsory education are presented as mean ± SD, range, and 95% CI. All other variables are shown as the number of participants, percentages in parentheses.

|  |  | Case Group |  | Control Group |  |
| --- | --- | --- | --- | --- | --- |
|  |  | Women<br>(n=12) | Men<br>(n=27) | Women<br>(n=270) | Men<br>(n=81) |
| Age (at the time of responding) | Mean ± SD | 33.8 ± 6.9 | 33.8 ± 7.5 | 38.7 ± 5.9 | 38.6 ± 5.9 |
|  | Range | 24–43 | 23–49 | 23–52 | 20–49 |
|  | 95%CI | [29.2, 38.3] | [30.8, 36.8] | [38.0, 39.4] | [37.3, 39.9] |
| Relation to the target child | Genetic parent | 11 (91.7%) | 14 (51.9%) | 269 (99.6%) | 80 (98.8%) |
|  | Step or adoptive parent | 0 (0%) | 6 (22.2%) | 1 (0.4%) | 1 (1.2%) |
|  | Cohabitant of the caregiver | 1 (8.3%) | 7 (25.9%) | 0 (0%) | 0 (0%) |
| Marital status (at the time of responding) | Unmarried | 1 (8.3%) | 2 (7.4%) | 2 (0.7%) | 1 (1.2%) |
|  | Legally married | 7 (58.3%) | 10 (37.0%) | 235 (87.0%) | 78 (96.3%) |
|  | Married (common-law) | 1 (8.3%) | 0 (0%) | 2 (0.7%) | 0 (0%) |
|  | Married (not specified) | 0 (0%) | 0 (0%) | 2 (0.7%) | 1 (1.2%) |
|  | Separated | 0 (0%) | 0 (0%) | 8 (3.0%) | 0 (0%) |
|  | Divorced or widowed | 2 (16.7%) | 15 (55.6%) | 21 (7.8%) | 1 (1.2%) |
|  | Unknown | 1 (8.3%) | 0 (0%) | 0 (0%) | 0 (0%) |
| Household of the target child (during the target period) | Living with both genetic parents | 5 (41.7%) | 13 (48.1%) | 237 (87.8%) | 79 (97.5%) |
|  | Single parent | 1 (8.3%) | 2 (7.4%) | 26 (9.6%) | 1 (1.2%) |
|  | Living with a non-kin caregiver | 6 (50.0%) | 12 (44.4%) | 6 (2.2%) | 1 (1.2%) |
|  | Other | 0 (0%) | 0 (0%) | 1 (0.4%) | 0 (0%) |
| Educational background | Junior-high school graduate & dropout | 7 (58.3%) | 15 (55.6%) | 5 (1.9%) | 0 (0%) |
|  | High school / vocational school (post-junior high school) graduate | 3 (25.0%) | 11 (40.7%) | 44 (16.3%) | 13 (16.0%) |
|  | University / junior college / vocational school (post-high school) graduate | 2 (16.7%) | 0 (0%) | 194 (71.9%) | 55 (67.9%) |
|  | Master's degree or higher | 0 (0%) | 1 (3.7%) | 26 (9.6%) | 13 (16.0%) |
|  | Unknown | 0 (0%) | 0 (0%) | 1 (0.4%) | 0 (0%) |
| Years of post-compulsory education | Mean ± SD | 2.5 ± 2.0 | 2.0 ± 1.9 | 5.8 ± 1.8 | 6.4 ± 1.8 |
|  | Range | 0–7 | 0–9 | 0–9 | 2–9 |
|  | 95%CI | [1.1, 3.8] | [1.3, 2.8] | [5.6, 6.0] | [6.0, 6.8] |
| Gender of the target child | Female | 6 (50.0%) | 7 (25.9%) | 114 (42.2%) | 36 (44.4%) |
|  | Male | 6 (50.0%) | 19 (70.4%) | 155 (57.4%) | 43 (53.1%) |
|  | Male & female twin | 0 (0%) | 0 (0%) | 1 (0.4%) | 1 (1.2%) |
|  | Unknown | 0 (0%) | 1 (3.7%) | 0 (0%) | 1 (1.2%) |
| Type of crime convicted | Homicide | 0 (0%) | 5 (18.5%) | .. | .. |
|  | Injury / Injury Causing Death | 7 (58.3%) | 17 (63.0%) | .. | .. |
|  | Unlawful Capture or Confinement Causing Death or Injury | 1 (8.3%) | 2 (7.4%) | .. | .. |
|  | Abandonment by a Person Responsible for Their Care / Abandonment Causing Death or Injury | 3 (25.0%) | 2 (7.4%) | .. | .. |
|  | Other | 1 (8.3%) | 1 (3.7%) | .. | .. |

### The Control group

To establish a comparable child-rearing population in Japan, we recruited 351 adults (270 women, 81 men) who had child-rearing experience during a period similar to that of the Case group. The birth years of the target children were aligned across the two groups (Control: 2013.7 ± 4.5 [95%CI 2013.3, 2014.2], Case: 2013.6 ± 4.3 [2012.2, 2015.0]). As a proof-of-concept study to comprehensively characterize the life-course factors underlying severe maltreatment, we did not attempt to match other parameters in order not to exclude the influence of key social-ecological variables (e.g., age, income). Rather, we used an Analysis of Covariance (ANCOVA) to control some of the key parameters (see below). We also verified the representativeness of our samples by comparing the Control group with national data from the Comprehensive Survey of Living Conditions ^20^ and the Population Census of Japan ^21^, and the Case group with those in the Child Maltreatment Fatality Reviews ^17^ and national statistics on newly imprisoned offenders ^22^ (Fig. S1).

There was a notable female bias in the control group, whereas the Case group comprised more males. To mitigate this bias, all statistical analyses were stratified by gender to ensure that gender-specific profiles were accurately captured.

### The relative impact of life-course factors on child maltreatment

Our survey comprised 402 quantitative or qualitative (open-ended) questionnaire items, retrospectively encompassing the family-of-origin (FoO) conditions, childhood adversity, experiences in compulsory schools, socioeconomic status, behavioral and cognitive traits, medical and neurobiological histories, marital and childcare experiences, conditions of their child(ren), household environments, and the stressors and reflection at the target period (i.e., at the time preceding maltreatment incidence for the Case group and at the comparable time for the Control group; see Methods). 70 numerical parameters (Fig. S2; hereafter *italicized*) were extracted and compared between genders in each group using Welch’s t-test. As numerous gender differences were observed in both groups, subsequent group comparisons were stratified by gender and conducted using analysis of variance (ANOVA) (Fig. 1, S2). Although the sample size of the Case group (women, n = 12; men, n = 27) was constrained by the availability of participants, a post-hoc power analysis indicated that the study had statistical power greater than 0.8 to detect a medium or large effect sizes (_W_^2^ >.06) at an alpha level of 0.05 (Fig. S2), confirming that the study was sufficiently powered to identify the key risk factors in our comparisons.

**Figure 1.**
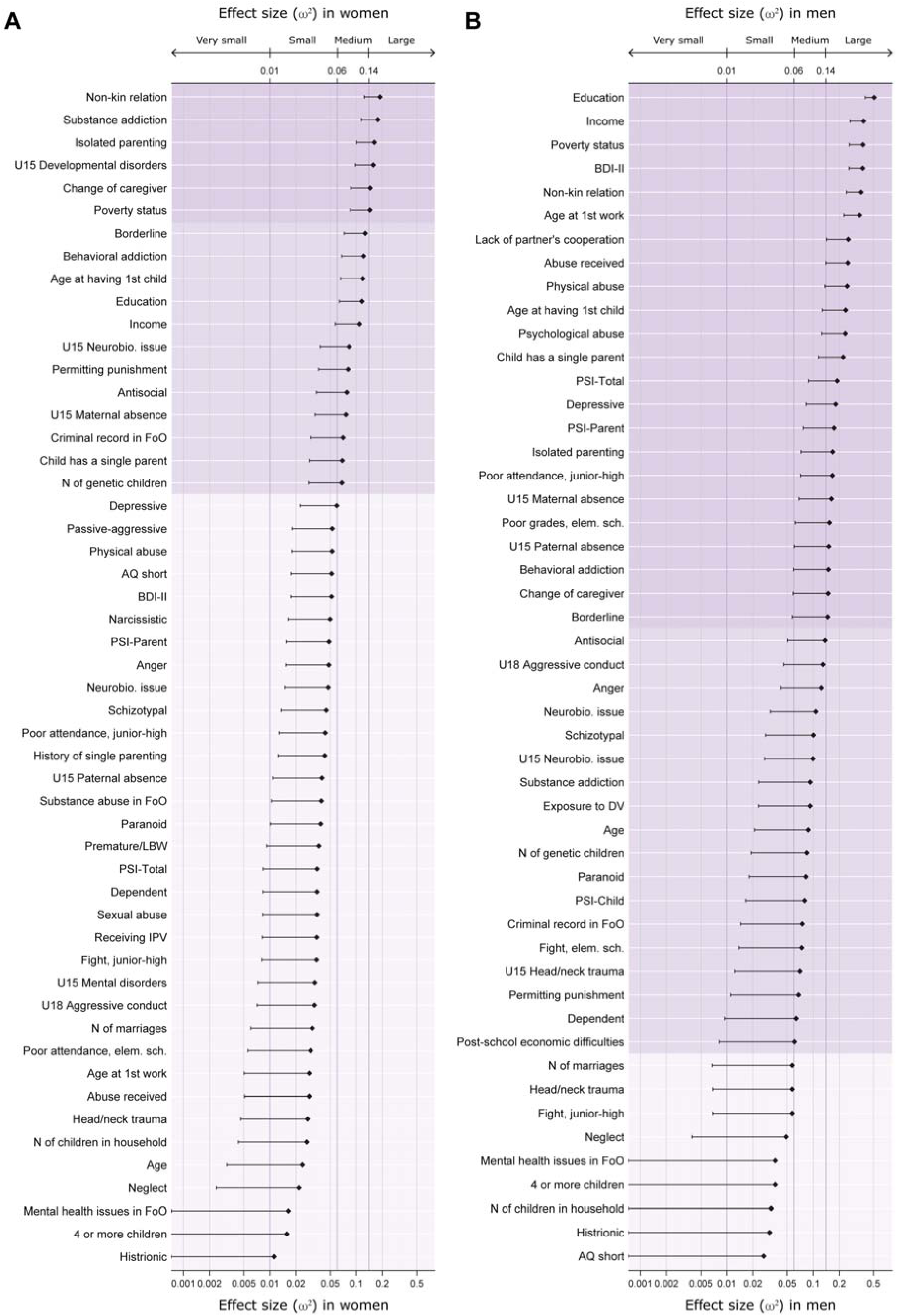
Standardized effect sizes of group differences across 73 background factors derived from ANOVA. The x-axes represent the effect sizes (omega-squared, _ω_^2^) of the Group main effect for women (**A**) and men (**B**), derived from ANOVA. The Group factor showed negative effects on all variables except *age* (*at the time of responding to the questionnaire*, *first working experience*, and *having the first child*), *education*, and *annual income*. For visibility, the x-axis is shown on a log scale. Background shading denotes the interpretation ranges for _ω_^2^ (very small < .01; small < .06; medium < .14; large ≥ .14). Error bars represent one-sided 95% confidence intervals. Note that several confidence intervals fall outside the displayed plot range; see Figure S2 for the precise values. AQ: Autism-Spectrum Quotient. BDI-II: Beck Depression Inventory, 2nd Edition. DV: domestic violence. Elm. sch.: elementary school. FoO: family of origin. IPV: intimate partner violence. LBW: low birth weight. N: number. PSI: Parenting Stress Index. U15: age 15 or younger. U18: age 18 or younger. *Neurobiological issue* scores were calculated from sensory impairment, diagnosed developmental and mental disorders, head-neck trauma with the loss of consciousness or medical treatment, and substance addiction. See Table S2 for the definition of 73 parameters analyzed.

The effect sizes (ES; _W_^2^) derived from the ANOVAs represent the raw magnitude of group differences (Fig. 1, S2). Large ESs (_W_^2^ ≥ .14, P < .05) were observed for three environmental-domain and one developmental-domain parameters in both genders:

▪ *Isolated parenting*, a lack of childcare support (w ES .161, one-sided 95% CI lower side [.101], m .167 [.073]).
▪ *Non-kin relation,* the presence of a non-genetic child in the household (w .188 [.124], m .358 [.241]).
▪ *Poverty status* (w .144 [.086], m .377 [.260]. Also see Fig. S1F).
▪ *Change of caregiver,* the number of primary-caregiver changes experienced by the participant by age 15 (ES: w .145 [.087], m .148 [.059]. Mean: Control: w 0.3, m 0.2 times, Case: w 1.8, m 0.9 times. Also see Fig. S3)

Additionally, 8 parameters showed a large ES in one gender and a medium ES in the other: *U15 maternal absence* (the duration during which the participant did not live with their genetic mother before age 15), *education* (years of post-compulsory education), *borderline-*like behavioral/cognitive trait, *substance addiction, behavioral addiction* (sex, gambling, shopping, and eating), *age of having 1^st^ child, single parenting* (the genetic parent of the target child being legally a single parent), and *income*.

As observed in incarcerated populations globally ^23^ (Fig. S1G), the Case group had significantly fewer years of *education*; on average, more than 3 years fewer for both genders (Table 2, Fig. 2A). They were also modestly younger at the time of the survey (hereafter *age*), with a mean difference of more than 4 years (Table 2, Fig. 2B). These findings are plausibly due to the fact that the Case participants had children significantly earlier (*age at having 1st child,* Control: w 30.5, m 31.9 y/o, Case: w 22.0, m 24.5 y/o. c.f. General population in Japan, 2015, w 30.7, m 32.8 ^24^). To assess whether the identified risks were independent of these demographic disparities, we subsequently conducted an Analysis of Covariance (ANCOVA) using *age* and *education* as covariates (Fig. 3, S2). Crucially, even after adjusting for these covariates, *isolated parenting* and *non-kin relations* continued to show large ESs in both genders. Similarly, parameters showing a large ES in one gender and a medium ES in the other included *U15 maternal absence, change of caregiver, substance addiction, behavioral addiction,* and *single parenting* (the genetic parent of the target child being legally a single parent). In contrast, the ES of *poverty status* decreased to medium, and the ESs for *age of having 1^st^ child* and *income* decreased to small (see Fig. S1F). These findings suggest the robust impact of immediate environmental factors, particularly isolated parenting and non-kinship, on the occurrence of severe child maltreatment, independent of the caregivers’ age or educational background.

**Figure 2.**
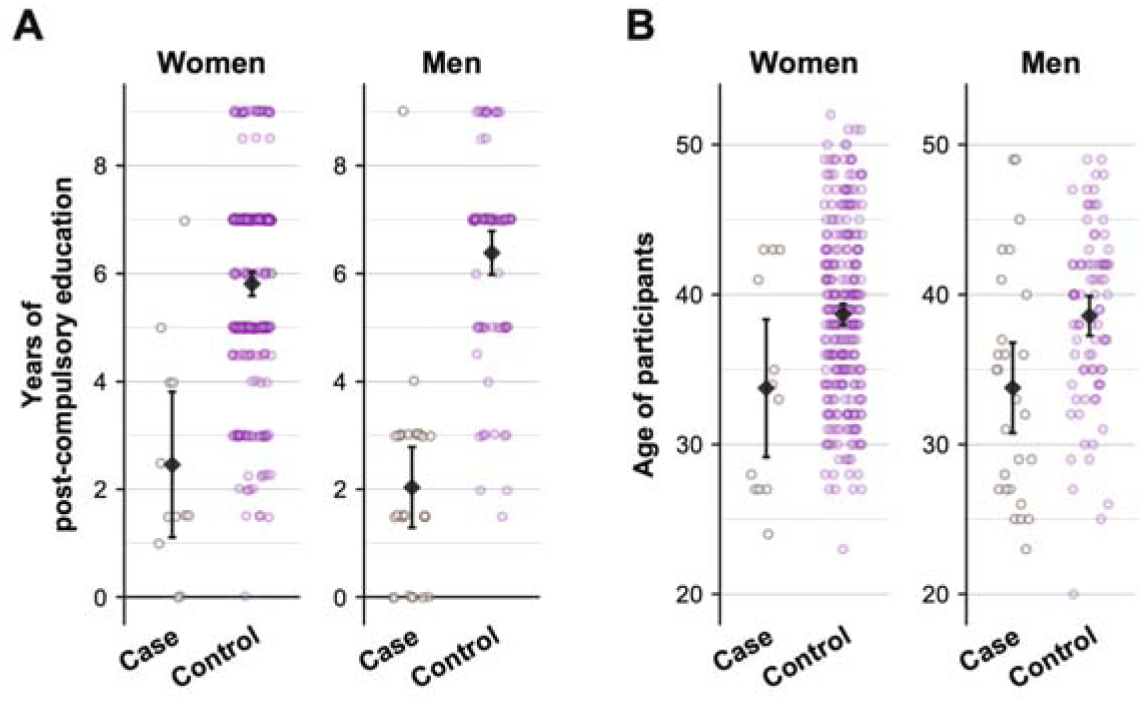
Age and educational attainment of participants. **A.** Age of the participants at responding to the questionnaire. There was a significant difference between the Case and Control groups (Welch’s t-test; women P = .039, Hedges’ *g* = -0.70 [-1.34, -0.03]; men P = .005, *g* = -0.69 [-1.16, -0.21]). **B.** The years of post-compulsory education of the participants. There was a significant difference between the Case and Control groups (women P < .001, *g* = -1.58 [-2.42, -0.71]; men P < .001, *g* = -2.30 [-2.94, -1.64]).

**Figure 3.**
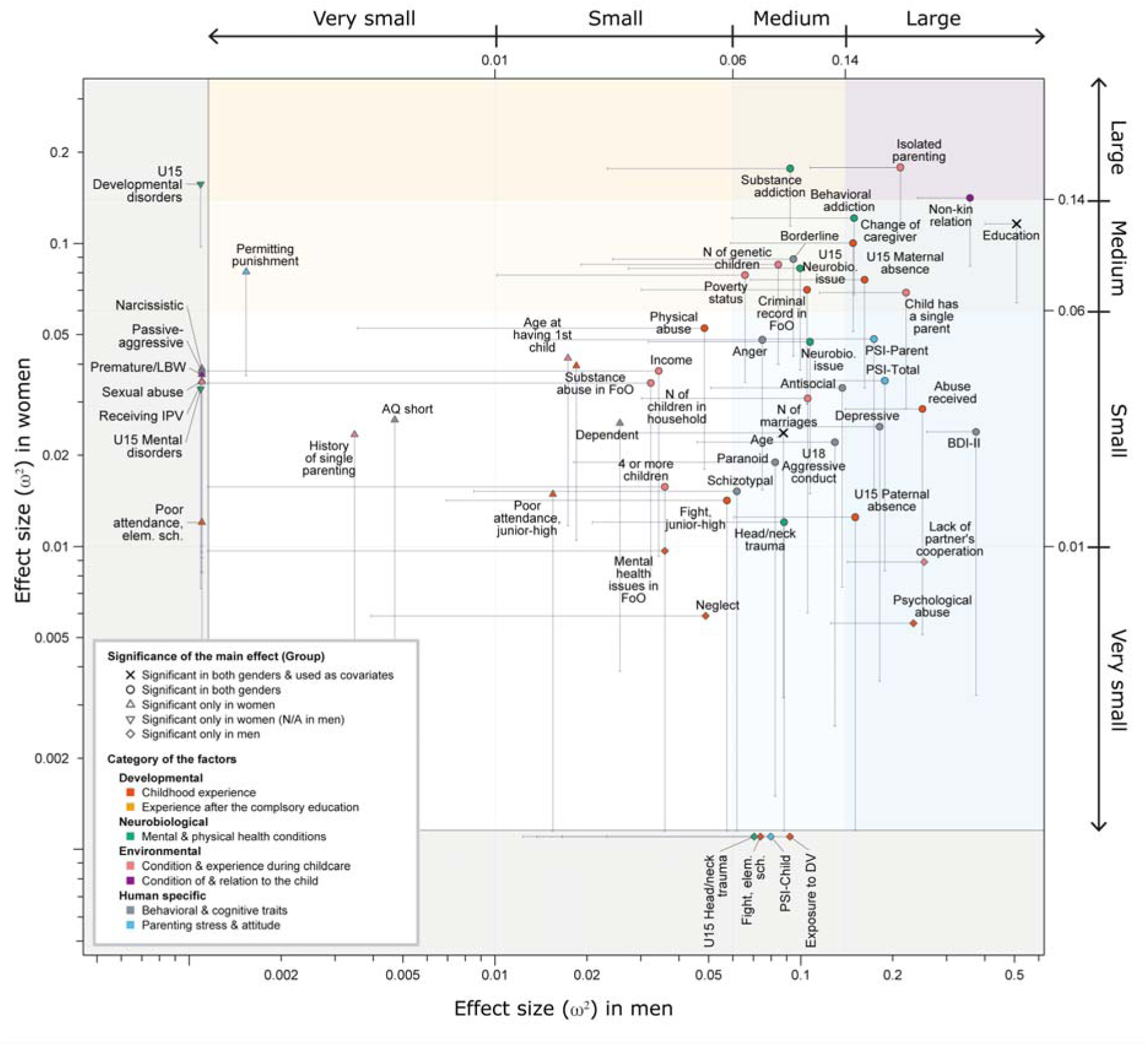
Standardized effect sizes of group differences across 73 background factors derived from ANCOVA. The x- and y-axes represent the effect sizes (omega-squared, _ω_^2^) of the Group main effect for men and women, respectively, derived from ANCOVA adjusted for *age* and *education*. The Group factor showed negative effects on all variables except *age* (*at the time of responding to the questionnaire*, *first working experience*, and *having the first child*), *education*, and *annual income*. Marker shapes indicate statistical significance. For visibility, both axes are shown on a log scale, and points with _ω_^2^ < .001 on either axis are aligned within the gray areas. Background shading denotes the interpretation ranges for _ω_^2^ (very small < .01; small < .06; medium < .14; large ≥ .14). Error bars represent one-sided 95% confidence intervals. Note that several confidence intervals fall outside the displayed plot range; see Figure S2 for the precise values. AQ: Autism-Spectrum Quotient. BDI-II: Beck Depression Inventory, Second Edition. DV: domestic violence. Elm. sch.: elementary school. FoO: family of origin. IPV: intimate partner violence. LBW: low birth weight. N: number. PSI: Parenting Stress Index. U15: age 15 or younger. U18: age 18 or younger. *Neurobiological issue* scores were calculated from sensory impairment, diagnosed developmental and mental disorders, head-neck trauma with the loss of consciousness or medical treatment, and substance addiction. See Table S2 for the definition of 73 parameters analyzed.

### Gender-specific effects of background factors

Our ANOVA and ANCOVA analyses revealed distinct vulnerability profiles between genders (Fig. 1, 3, S2), thus the ANCOVA results are described here: A large ES was observed for *U15 paternal absence* in men (.151 [.061]), whereas the effect was small in women (.012 [.000]), suggesting a specific impact of childhood paternal presence on future fatherhood.

ESs of depressive tendency, assessed using BDI-II (Beck Depression Inventory, past two weeks) and SCID-II (Structured Clinical Interview for DSM-IV Axis II Personality Disorders; past several years) ^25,26^ as well as the parental domain of the *Parenting Stress Index* (PSI) ^27^ were large only in men. These measures were significantly intercorrelated (BDI-II vs. SCID-II Depressive: r = 0.66 [0.61, 0.72], BDI-II vs. PSI-Total: r = 0.55 [0.47, 0.61], SCID-II Depressive vs. PSI-Total: r = 0.50 [0.42, 0.57]), Similarly, total *abuse received* (comprising physical, psychological, and sexual abuse, neglect, and exposure to domestic violence) and *psychological abuse* received, which were strongly correlated (*r* = .87 [.85, .90]), showed large ESs only in men (.234 [125]; .250 [.140]). This result may be attributed to the higher baseline scores of depressive tendencies and reported abuse among women than men in the Control group, consistent with previous literature ^28–30^.

Conversely, the participants’ *developmental disorders diagnosed by age 15* showed a large ES uniquely in women, although this was based on only two Case women (one attention-deficit/hyperactivity disorder (ADHD) and one Asperger’s). These results suggest that although certain stressors are universal, the extent to which they differentiate maltreating from non-maltreating individuals is gender-dependent.

### Parenting-inhibitory factors across mammals are accumulated in child maltreatment convicts

Next, to assess the prevalence and cumulativity of cross-species maltreatment risks, 24 key parameters were selected as human counterparts to established risk factors for mammalian parenting across five domains ^12^ (Table 1): (i) developmental (early adversities, including childhood parental absence, received physical abuse, and neglect; premature parenthood); (ii) neurobiological (e.g., sensory impairment, head trauma, mental disorders, substance addiction); and (iii) environmental, further subdivided into (iii-1) non-kin relation, (iii-2) child factors (e.g. multiple birth, multiple children, child’s health issues), (iii-3) socio-ecological hardships during parenthood (e.g., poverty, isolated or single parenting, receiving intimate partner violence) (Table 1). Each factor was binary or binarized using a threshold of the Control group mean +2SD (within gender).

Analyses of these 24 parameters showed that the Case participants accumulated more risk factors than Controls, representing a 4.62-fold increase for women and a 5.87-fold increase for men (Table 1, Fig. S4). When human-specific maltreatment types (*exposure to DV*, *psychological abuse*, and *sexual abuse*) were included, the disparities remained significant (4.43- and 5.76-fold increases for women and men, respectively; Table 1, Fig. S4).

To visualize the relative prevalence (Fig. 4A, B, D, E) and odds ratios (Fig. 4C, F) of the five risk domains, we coded each participant as positive for a given domain (i–iii) if they possessed at least one parameter within that domain. We then quantified the number of participants positive for each domain (i–iii) in the left Venn diagram of Fig. 4, and for the environmental subdivisions (iii-1 to iii-3) in the right Venn diagram. Among Controls, 33.3 % of women and 49.4 % of men were negative for all of (i-iii) domains, while 100 % of Case participants were positive for at least one domain. The prevalence of most risk domains was significantly higher in the Case group relative to the Control group for both genders, except for the neurobiological factors (ii) in women and for the child factors (iii-2) in men (Table 1, Fig. 4C, F). The accumulation of risks across multiple domains was more prevalent in the Case group; 25.0 % of women and 40.7 % of men in the Case group exhibited risks in all three domains (i-iii) simultaneously, compared to only 5.6% and 1.2 % in the Control group. The overlapping area between (i) and (iii) are significantly different between groups. Furthermore, 25.0 % of women and 11.1 % of men in the Case group had risks across all three environmental subdomains, whereas this was not observed in any Control participants. The results were similar when human-specific developmental factors (*exposure to DV*, *psychological abuse*, and *sexual abuse*) were included (Fig. 4, numbers in parentheses; Fig. S4). Together, these findings suggest that in contemporary Japanese society, evolutionarily conserved mechanisms underlying parental motivation loss remain highly relevant, yet precipitate into severe maltreatment only when there is substantial multisystemic accumulation of the stressors.

**Figure 4.**
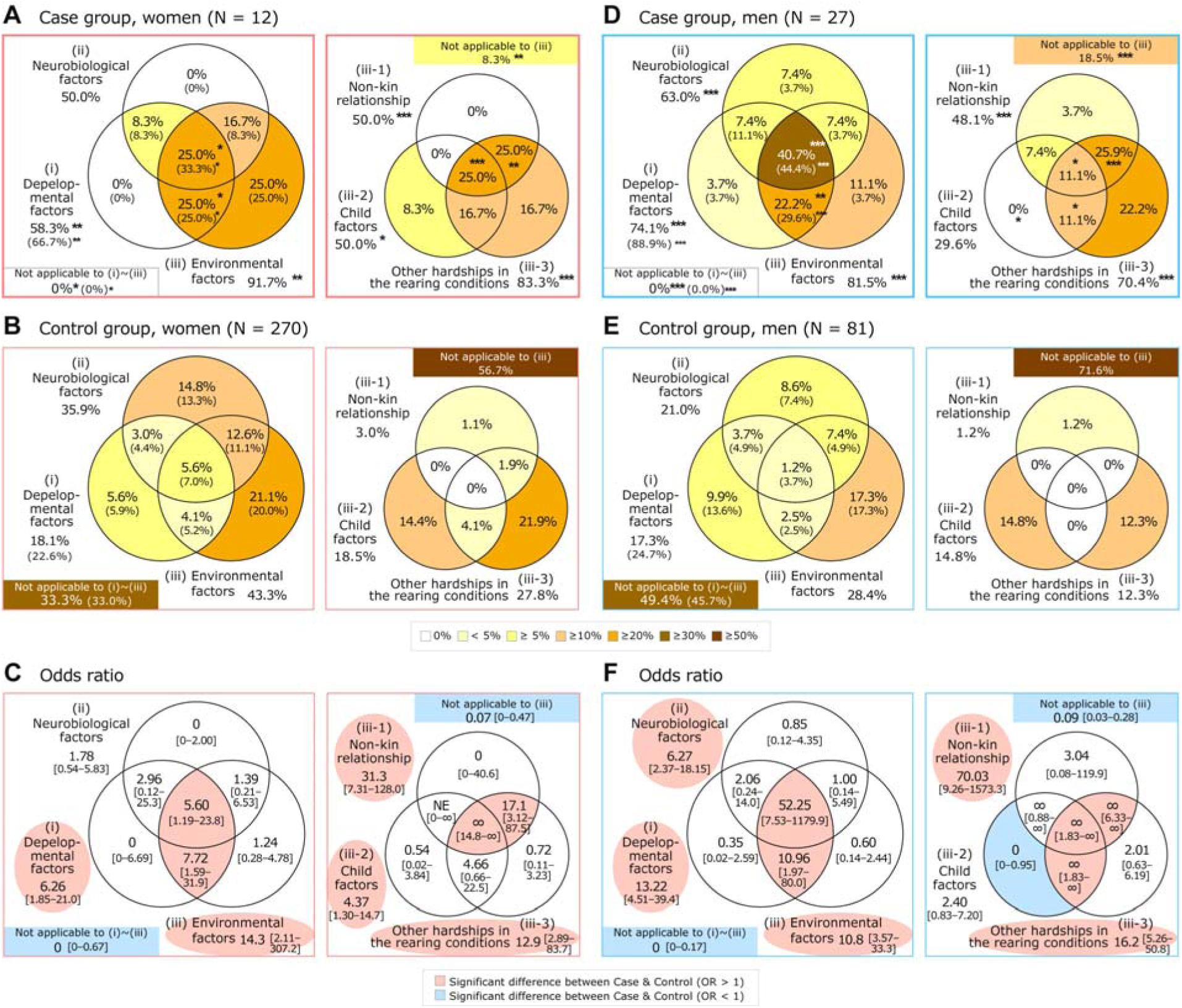
Prevalence and co-occurrences of the developmental, neurobiological, and environmental factors conserved across mammals. **A, B, D, E.** The Venn diagrams show the percentage of subjects who possess at least one of the human counterparts of the parenting-inhibitory factors associated with offspring desertion/attack in non-human mammals ^12^. The factors are divided into three domains (i–iii), and the domain (iii) is further subdivided as described in Table 1. The percentages in the parentheses for domain (i) include human-specific early adversity, namely sexual abuse, psychological abuse, and exposure to domestic violence. Asterisks denote statistical significance based on Fisher’s exact test comparing the Case and Control groups (* P < .05, ** P < .01, *** P < .001). **C, F**. Odds ratios and 95% confidence intervals for each area of the Venn diagrams, with the Control used as the reference.

### Potential developmental pathways to severe child maltreatment

The multiple risk factors during parenthood among Case participants may not accumulate randomly; rather, they appear to be the result of a snowball effect originating in early-life adversities ^6,31^. To gain insight into the relationship between early-life factors and later maltreatment, we constructed a developmental pathway model using structural equation modeling (SEM) ^32^ (see ^9^ for the caveats of this approach). We selected 17 key parameters in Fig. 5 based on their significance in Fig. 1 and 3, relevance in mammalian models, and prevalence in existing human literature, while excluding redundant variables; (e.g., the *number of primary-caregiver changes by age 15* was excluded due to high collinearity with the *U15 maternal/paternal absence*). These 17 parameters were sorted into four temporal layers: childhood, youth, immediate household environment and parenting attitude (Fig. 5A). The SEM estimated standardized total effects (STE) on group (Case vs. Control) of each parameter in the specified model, presuming the remainder of the model remains constant. STEs represent combined direct and indirect associations along temporally-ordered paths of each parameter with the occurrence of severe maltreatment (i.e., temporal relatedness, not causality) (Fig. S5A, genders combined; Fig. S5B, women; Fig. S5C, men).

**Figure 5.**
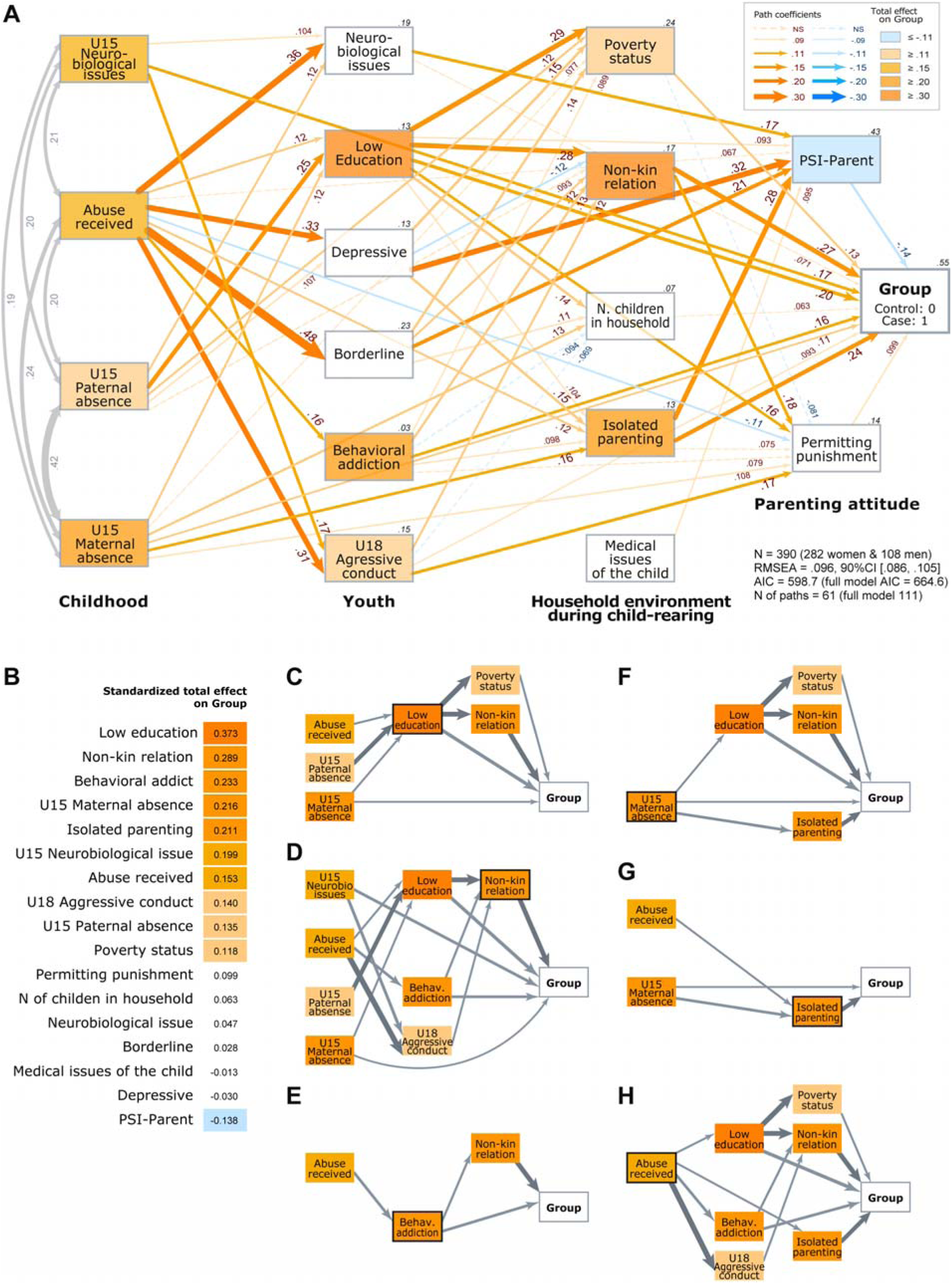
Developmental pathway model of the major risk factors. **A.** The final SEM model. Rectangles represent observed variables, and the adjusted R^2^ value is displayed in the top right corner. The color of each rectangle indicates its standardized total effect on ‘Group’. One-handed straight arrows represent standardized partial regression coefficients, and two-handed curved arrows represent correlation coefficients. Arrow widths are proportional to the absolute values of the coefficients (0.10 = 1pt). Dashed arrows indicate paths retained in the final model but with P > .05. N: number. PSI: Parenting Stress Index. U15: age 15 or younger. *Neurobiological issue* scores were calculated from sensory impairment, diagnosed developmental and mental disorders, head-neck trauma with the loss of consciousness or medical treatment, and substance addiction. *Low education* was defined as the min–max–normalized years of post[compulsory education (maximum of 9 years), with the resulting score reverse[scored so that lower education levels corresponded to higher values. *Depressive* score was based on the SCID-II. See Table S2 for the definition of parameters. **B.** Standardized total effects on ‘Group,’ presented in descending order. Cell colors correspond to the legend in panel A. **C–H**. Extracted nodes and pathways highlighting the effects of low education (C), non-kin relation (D), behavioral addiction (E), U15 maternal absence (F), isolated parenting (G), and abuse received (H). The following nodes and paths were removed: nodes with standardized total effect on ‘Group’ below 0.11, paths with coefficients below 0.11.

Positive STEs were largest for the following parameters in descending order: *low education, non-kin relation, behavioral addiction, U15 maternal absence, isolated parenting, U15 neurobiological issues*, *abuse received*, *U18 aggressive conduct, U15 paternal absence,* and *poverty status* (Fig. 5B) (for visualization, educational attainment in Fig. 1 and 3 was reverse-scored to represent “low education” in Fig. 5B so that all parameters have higher means in the Case group). To summarize the complex temporal relations, we removed 7 factors with STEs < 0.1 from Fig. 5A, and then drew summary diagrams for each of the top 5 factors plus *abuse received*, retaining only paths that traverse the focal factor or have path coefficients ≥ 0.1. (Fig. 5C-H). These diagrams suggest that caregiving breakdown pathways can be coarsely categorized as follows:

1. The Socio-Economic Path. Childhood absence of genetic parents (particularly fathers, possibly due to the gender wage gap) (Fig. 3C), preceded low educational attainment. This, in turn, was associated with severe maltreatment (Group) both directly and indirectly via poverty and non-kin caregiving relationships (Fig. 5D). The connection between low education and non-kin relations may be mediated by the earlier age of having the first child, which significantly correlates both with education (w: r = .39 [.28, .48], m: .41 [.24, .56]) and non-kin relation in the family (w: -.27 [-.38, -.15], m: -.48 [.61, -.32]).
2. The Neuro-Behavioral Path. *U15 neurobiological issues* (head/neck trauma and developmental disorders) and *abuse received* (Fig. 5D, E, G, H) correlated with externalizing behavior (*U18 aggressive conduct, behavioral addiction*), which preceded severe child maltreatment directly or indirectly through *isolated parenting*. *U15 maternal absence* (Fig. 5F) precedes Group both directly and via *isolated parenting*. *U15 neurobiological issues* also showed a significant direct effect.

Notably, the STE of *PSI-parent* (parenting stress) was negatively correlated with education and with Group when other background risks were controlled, despite higher raw PSI scores in the Case group. This suggests that the ability to perceive and report parental stress may function as a resilience factor. Additionally, the STEs of *the number of children in households* and *the medical issues of the child* were not significant. This suggests that while the excessive demands arising from child-related factors are often critical drivers of caregiving failure in non-human mammals, their effects may be ameliorated in modern human society.

As complex life-course models may be unstable especially when the sample size was small ^9^, we also conducted gender-stratified modeling (Fig. S5) and confirmed that gender differences in STEs are expectable from the result of Fig. 1. The gender-specific models also demonstrated that the STEs for the absence of the same-sex parent were significant, whereas those for the opposite-sex parent were not, suggesting that the presence of the same sex genetic parent during childhood may act as a protective factor. While early adversity and educational barriers were more dominant among men, environmental constraints (e.g., isolation, non-kinship) exerted a larger total effect among women, mirroring our earlier findings (Fig. 3). These data suggest that interventions should be tailored to these gender-specific developmental vulnerabilities.

## Discussion

### Major findings

This study quantitatively demonstrates that severe child maltreatment in a modern society exhibits shared risk structures with caregiving breakdown in non-human mammals, most prominently isolated parenting and non-kinship (Fig. 1, 3, 4). In social mammals that care infants cooperatively, infant survival depends on a network of “alloparents” or helpers ^33–35^. Also in traditional human societies, infant allocare by maternal grandmothers or siblings has been a near-universal practice and significantly enhances infant survival ^36,37^. Consistently, our data identified that maternal absence in childhood is associated with *isolated parenting*, which is the most critical antecedent of maltreatment among women, plausibly due to the lack of postpartum care by maternal grandmother, *Satogaeri shussan,* in Japanese tradition.

This study has also demonstrated that the standardized total effect of the non-kin relationship on severe maltreatment is among the top three for both genders (Fig. S5). In Japan, the prevalence of stepfamilies was reported as 2.1 % among 2258 responded mothers at 4-month infant checkup ^38^, which is comparable to our Control group (w 2.2%, m 1.2%). It may be underestimation, as 8.4 % of all couple families with children of any age were stepfamilies in Canada ^39^, and that 6.3-7.5 % of children aged 17 and under are living with a stepparent in New Zealand ^40^. Nevertheless, the prevalence of non-kin relations in the Case group (w 50.0 %, m 44.0 %) is remarkably high, even compared with the CMFR cases (10.5%, Fig. S1D). This data together with existing human literature ^15,41–44^ apparently aligns with studies of non-human mammals ^13,45^.

However, it should also be emphasized that neither isolated parenting nor non-kin relationship appeared as a solitary risk factor among Case participants (Fig. 4); e.g., one Case man representing (iii-1) had other background factors in (i) and (ii). In other words, when these proximal conditions are not preceded by other adversities, they do not associate with severe maltreatment as in Control participants. Thus, our data align with Belsky’s argument that the severe maltreatment of non-kin children may not be “adaptive” in modern societies unless the environmental conditions are impoverished ^14,46^. These observations instead highlight the importance of societal support for stepfamilies experiencing multiple hardships.

Among human-specific factors, educational attainment is a notable antecedent of severe child maltreatment, particularly among men. More than half of the Case participants did not graduate from high school. However, they all completed this 402-question survey written with *Kanji* (Chinese characters) without *Furigana* (reading aid), which required substantial reading & writing skills and attention. Thus, their low educational attainment appears disproportionate to their intellectual level and may be due to the families’ functional limitation to support their education. Of relevance, a prospective cohort study in the UK found associations between ACEs and lower educational attainment that remained after adjusting for family and socioeconomic factors ^47^. In such cases, low educational attainment should be viewed as an index of early adversity, rather than a neurobiological factor or an individual choice.

These data may suggest two foci of social work in preventing child abuse in Japan: educational support to children in adverse family environments, and parenting support to families experiencing multiple hardships including complex family relationships and isolated parenting. However, several limitations should be considered when interpreting the present data, as described in the next section.

### Limitations

This study compares (near-)lethal maltreatment in humans and animals by utilizing data from a rare incarcerated population, an approach that inherently involves several caveats. To fully leverage this unique dataset, we have conducted a comprehensive range of analyses; however, the following limitations must be carefully considered when interpreting the findings.

First, our retrospective case-control study identifies associations rather than causality. While a prospective cohort design is ideal for establishing causal relationships, the low prevalence of severe maltreatment makes such an approach impractical. Furthermore, although previous research has established the significant contributions of both genetic and environmental factors to the link between early adversities and later behaviors ^48–50^, direct biological investigations are challenging within incarcerated populations. Yet, while genetic predispositions are not targets for social interventions, this proof-of-concept study aims to present a testable hypothesis for future intervention research: implementing educational support for children in adversity and parenting support to multi-risk families, and evaluate the effect of such interventions. Nevertheless, integrating neuroimaging and genetic analyses in future studies remains essential to further validate the complex interplay between these biological and environmental risk structures.

Second, the sample size of Case participants (n=39) was constrained by the scarcity of incarcerated populations. Considering that the average annual number of child abuse deaths in Japan (CMFR cases) is 48.9 ± SD 6.78 (excluding murder-suicide, 2008-2018), and that the names of perpetrators are uncommonly published in the mass media (as far as we could ascertain, in 76 cases from 2013 to 2017, representing 33.6% of CMFR cases), our sample represents a substantial proportion of these rare, severe maltreatment cases. To expand this research, further initiatives led by the Japanese Ministry of Justice will be essential.

Third, our data relied on self-reported information. To enhance reliability, we cross-validated Case participants’ reports with public records and media outlets wherever available. We also implemented internal consistency checks by asking similar questions in different questionnaires a few weeks apart. Major discrepancies were rare, except that one Case man with grandiose tendency reported high income and educational attainment, which did not match with public records. The exclusion of his data did not impact the major findings of this study.

Finally, the present design may overlook the psychological diversity among perpetrators, such as variations in remorse or emotional detachment, which we intend to address in subsequent studies.

### Concluding remarks

Lastly, three remarks are discussed: First, no form of child maltreatment is justified in modern society, regardless of whether similar behaviors are observed in non-human mammals or are considered “evolutionarily adaptive” in certain biological contexts ^12,51^. The objectives of animal evolution – maximizing reproductive success – diverge fundamentally from the core values of modern society: the protection of human rights and child well-being. Nevertheless, even in contemporary societies, circumstances that threaten parents’ own survival inevitably constrain their capacity to nurture their young, and parental behaviors are governed by neurobiological systems that can be impaired by various causes.

Second, the present results align with the principle that group-level risk profiles cannot be generalized to individuals ^52^, as no single risk factor deterministically leads to child maltreatment. Especially non-kin relationships should not be stigmatized, yet professionals should be informed of the associated risks at the group level.

Third, risk factors should not be viewed as individual failures but as indicators for societal intervention ^53,54^. Even when household difficulties appear to be derived solely from parents’ maladaptive behaviors, such behaviors may be influenced by early adversities, and it is the children who will suffer the consequences unless adequate support is provided.

Interpreting the present findings with these remarks may facilitate the implementation of evolution-informed and balanced interventions to address contemporary child maltreatment.

## Acknowledgments

We extend our sincere gratitude to all the participants in this study, despite the difficult circumstances, and the Adult Correctional Services Division of the Correctional Bureau of the Ministry of Justice, the Legal Research Institute, and the penal institutions for their cooperation and advice, Kana Ohira and Kanako Nakamura for technical assistance, and Dr. Takeo Fujiwara for fruitful discussion.

## Author contributions

Y. S. and K.O.K. designed and carried out the survey. E.M. and Y.S. analyzed the data and produced the tables and figures. K.O.K. conceived of and organized the study. E.M., Y.S., and K.O.K. wrote the manuscript.

## Declaration of generative AI and AI-assisted technologies in the writing process

During the preparation of this work the author(s) used Grammarly, DeepL, Gemini, and Copilot, in order to improve the readability and language of the manuscript. After using this tool/service, the author(s) reviewed and edited the content as needed and take(s) full responsibility for the content of the published article.

## Data and code Availability

All data needed to evaluate the conclusions in the paper are present in the paper and the Supplementary Data. The raw data files can be provided by K.O.K. pending a material transfer agreement, due to ethical restrictions. No code is created in this study.

## Methods

### Participants

All experiments were approved by the RIKEN Wako Research Ethics Committee III (General research involving human subjects) and Ethical Committee on Research Involving Human Subjects of the Institute of Science Tokyo, and were conducted following the Ethical Guidelines for Medical and Biological Research Involving Human Subjects ^55^ and in accordance with the Declaration of Helsinki. Given the involvement of incarcerated individuals—a vulnerable population—special measures were taken to ensure voluntary participation and safety of the participants. Before conducting the research, we contacted the Adult Correctional Services Division of the Correction Bureau, Ministry of Justice, and its 62 correctional facilities, excluding those for juveniles, traffic offenders, and those with medical needs, to explain our research protocol and request permission for the survey. Three prisons (one women’s and two men’s) that declined were thus excluded from the study. Potential participants were informed in writing that their decision to participate, refuse, or withdraw would have no impact on their legal status or prison treatment. Written informed consent was obtained from all participants. To protect privacy, all data were pseudonymized immediately after collection and stored on a secure server with restricted access. For further validity and ethical aspects of this study, refer to ^56^.

### The Case group

By searching the database of major Japanese newspapers for child abuse cases reported from 2006 to 2018 (it generally takes 1 to 2 years to get the final sentence and the convict to be sent to prison after the occurrence of the incident), we identified the 302 convicts whose names and sentences they received were publicized. We sent written research explanations and invitation letters by postal mail to the 139 of these convicts who were presumed to be still imprisoned at that time. Out of the 86 convicts we were able to reach, 48 were interested in participating (55.8%). 81.3 % (39/48) of once-agreed participants could have finished all the questionnaires and been included in this study.

### The Control group

226 adults (152 women, 74 men) with experience in raising children were recruited through internet advertisements and free community papers and completed most of the questionnaires. In the latter part of this study, we specifically recruited men to compensate for the female bias of this group. Additionally, we recruited 125 parents (118 women, 7 men) who voluntarily attended a parenting support program, such as Parent-Child Interaction Therapy (PCIT) ^57^, CARE ^58^, and Triple-P ^59^. Although some of them have experienced mild difficulties in child rearing, none of them have been convicted of child maltreatment. Thus, we combined participants from these sources of recruitment as the Control group.

### Survey procedure

The survey questionnaire is presented as Table S1 Briefly, it was divided into four parts. Part 1 consisted of inquiries about demographic information, current and past marital/financial status, medical history, the composition of their family of origin and current family, educational history, experiences at elementary and junior-high schools, work history, experiences of traumatic events, and questions to measure their behavioral and cognitive traits (including Structured Clinical Interview for DSM-IV Axis II Personality Disorders (SCID-II) ^26^ and the Dark Triad ^60,61^), and Japanese version of Beck Depression Inventory, Second Edition (BDI-II) ^25^. Part 2 inquired about the history of the family of origin and adverse childhood experiences using the WHO’s Adverse Childhood Experiences International Questionnaire, ACE-IQ ^62^, and attachment style questionnaire and related questions ^63^. Part 3 asked about their childcare experiences, including the Japanese version of Parenting Stress Index Short Form ^27,64^ and a part of Parental Abusive Attitude Inventory (PAAI) ^65^. Part 4 asked about the childcare circumstances including their interactions with family, friends, and community services. For case participants, the focus of Part 4 was on the time when child maltreatment occurred. For control participants, the focus time was on the target childcare period described in the one after next paragraph.

To the Case participants who showed interest in participating in this study, we first sent the consent form and Part 1 of the questionnaires. Upon their return, we sent Part 2 together with the additional questions about the ambiguities in Part 1. Upon their return, we sent Parts 3 and 4 with additional questions, if any. To avoid traumatic reactions in participants when they recall incidents of child maltreatment, a special sheet covered Part 4. The sheet informed participants that this section contained questions about the maltreatment incident and advised them not to open it if recalling the incident might upset them. All the Case participants who had answered Parts 1_–_3 also answered Part 4. No trouble, such as manifestations of traumatic reaction, has been reported from the participants or their penal institutions. The average age of the victimized children in the Case group was 3.0 ± 3.0 (0 _–_14) y/o. The period between the time of the incident and the time of participating in this survey was 3.4 ± 2.1 years on average (range 0–8).

To the Control participants, we first sent the consent form and Part 1. Upon their return, we sent Parts 2-4 (questions in Part 4 asked about their general experiences). For their child-rearing experiences, the 125 Control parents who attended a parenting support program were instructed to write about the child for whom they participated in the program (mean child age 6.7 ± 3.0 y/o, 0–17). The other 226 Control parents were instructed to write about their youngest child (designated as “the target child”). Among 226, the 142 participants who had their youngest child as a preschooler (2.7 ± 1.7, 0–6) were instructed to answer questions regarding their current child-rearing experiences. The other 84 participants whose youngest child was older than 6 y/o (11.6 ± 3.8, 6–27) were instructed to answer the questions by recalling the time when their youngest child was a preschooler (designated as “the target period”). This design allowed us to evaluate the effect of answering about past or present child rearing, as all the Case participants answered about the past.

### Statistical Analyses

Statistical software IBM SPSS Amos 28 was used for structural equation modeling (SEM). All other statistical analyses were conducted using R 4.4.3 ^66^. The following R packages were used: *car* 3.1.3 for Type III tests ^67^; *effectsize* 1.0.0 for calculating and interpreting effect sizes ^68^; *exact2×2* 1.6.9 for performing Fisher’s exact test and estimating odds ratios using the minimum likelihood method ^69^; and *pwr* 1.3.0 for power analysis ^70^. Adobe Illustrator 29.4 was used to prepare the Venn and path diagrams.

### Measures

See Tables 1 & S2 for details of the measures and variables used in this study. Yes-no responses were converted into dummy variables (‘no’ = 0 and ‘yes’ = 1). Likert and other ordinal scales were min-max normalized [0, 1], unless otherwise noted.

### Group comparisons (**Fig. 1**, S2)

Each parameter was compared between the Case and Control groups using ANOVA with Group as the main effect, followed by analyses of covariance (ANCOVA) including two covariates: *years of post-compulsory education* and *age at responding to the questionnaire*. The significance of each explanatory variable was assessed using Type III F-tests, and the effect sizes were quantified using omega-squared (_ω_^2^). If either covariate was nonsignificant, it was removed from the model and ANCOVA was re-performed (or ANOVA was used if both covariates were nonsignificant). Gender differences within each group were examined using Welch’s t-test, with Hedge’s *g* reported as the effect size. Interpretation of _ω_^2^ and Hedges’ *g* followed Field ^71^ and Cohen ^72^, respectively, as implemented in the *effectsize* package (very small: _ω_^2^ < .01, g < .20; small: _ω_^2^ < .06, g < .50; medium: _ω_^2^ < .14, g < .80; large: _ω_^2^ ≥ .14, g ≥ .80). Binary variables were treated as continuous variables for ANOVA/ANCOVA so that the same type of effect size could be used for comparison across all variables. Results from generalized linear models (GLMs) with logistic regression applied to binary variables did not differ meaningfully from those reported in this paper.

### Venn diagram analysis (VDA) (**Fig. 2**)

For the VDA, background factors across the participants’ life histories were classified into three domains: (i) developmental, (ii) neurobiological, and (iii) environmental, with the environmental domain further subdivided into three subdomains, as described in Table 1 ^12^.

### Structural equation modeling (SEM)

The developmental pathway models (Fig. 5, S5) were constructed using SEM with a full information maximum likelihood estimation (FIML). The proposed model includes 18 variables representing five stages along the participants’ life history: 1) childhood (age 15 or younger), 2) youth, 3) household environment during childcare, 4) parenting attitude, and 5) parenting outcome (Control vs Case group).

We first specified a full model that included all possible paths from one stage to the subsequent stage, except that no paths from earlier stages to the *child’s medical issues* were specified. This full model was constructed separately for three datasets: both genders combined, women only, and men only. Next, for each dataset, we generated candidate models by removing paths whose standardized partial regression coefficients fell below progressively increasing thresholds (< .05, < .06, < .07, < .08, < .09, < .10, < .11), in order to obtain models that were both explanatory and parsimonious. Final models were selected based on the lowest AIC value: the < .07 threshold for the combined-gender dataset, < .08 for the women-only dataset, and < .10 for the men-only dataset, because AIC began to increase when the next threshold was applied.

## Supplementary Figures and Tables

**Table S1 Questionnaire used in this study.**

See the attached Excel file.

**Table S2 Variables used for Figures 1** and **3**.

See the attached Excel file.

**Figure S1.**
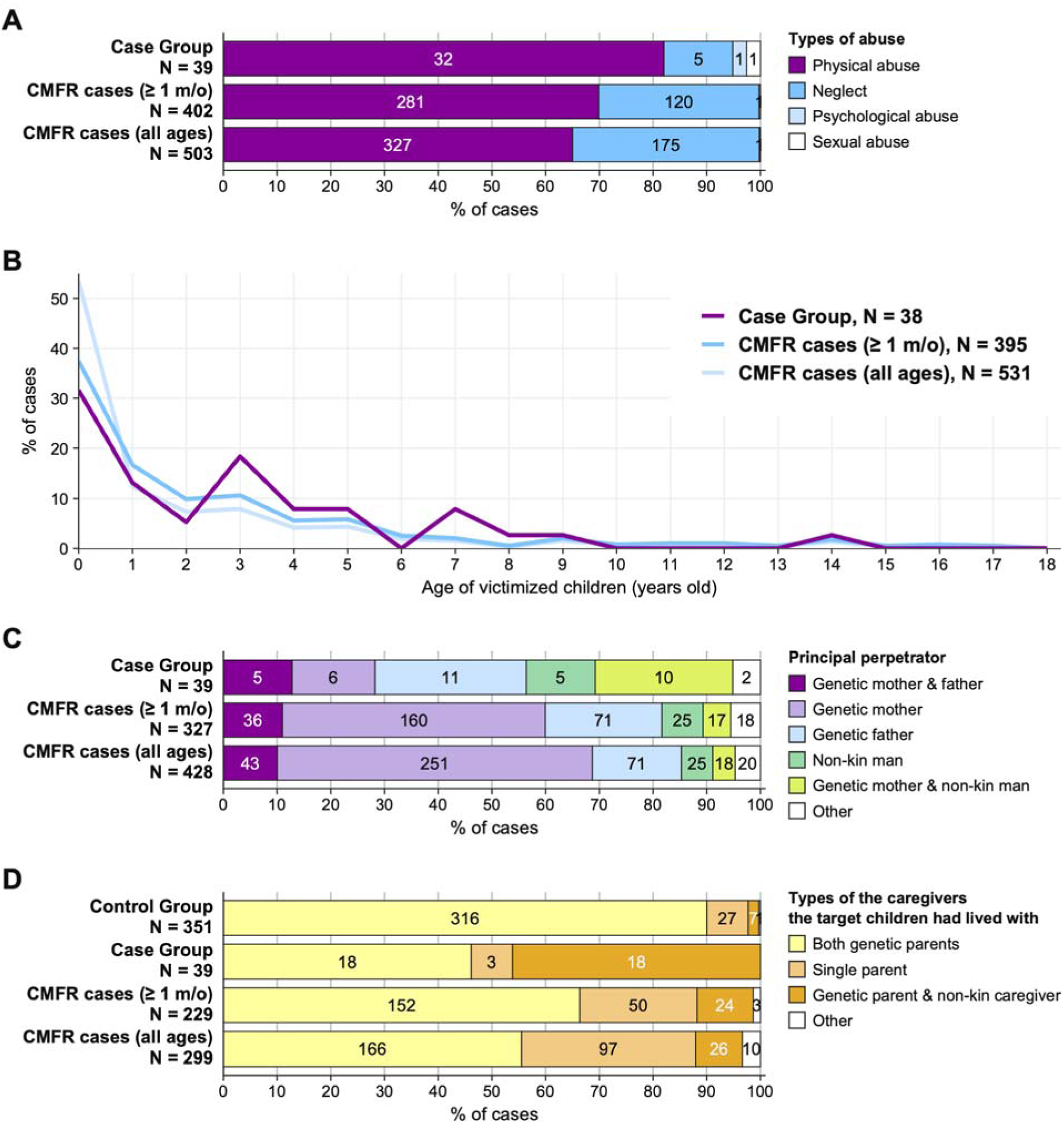
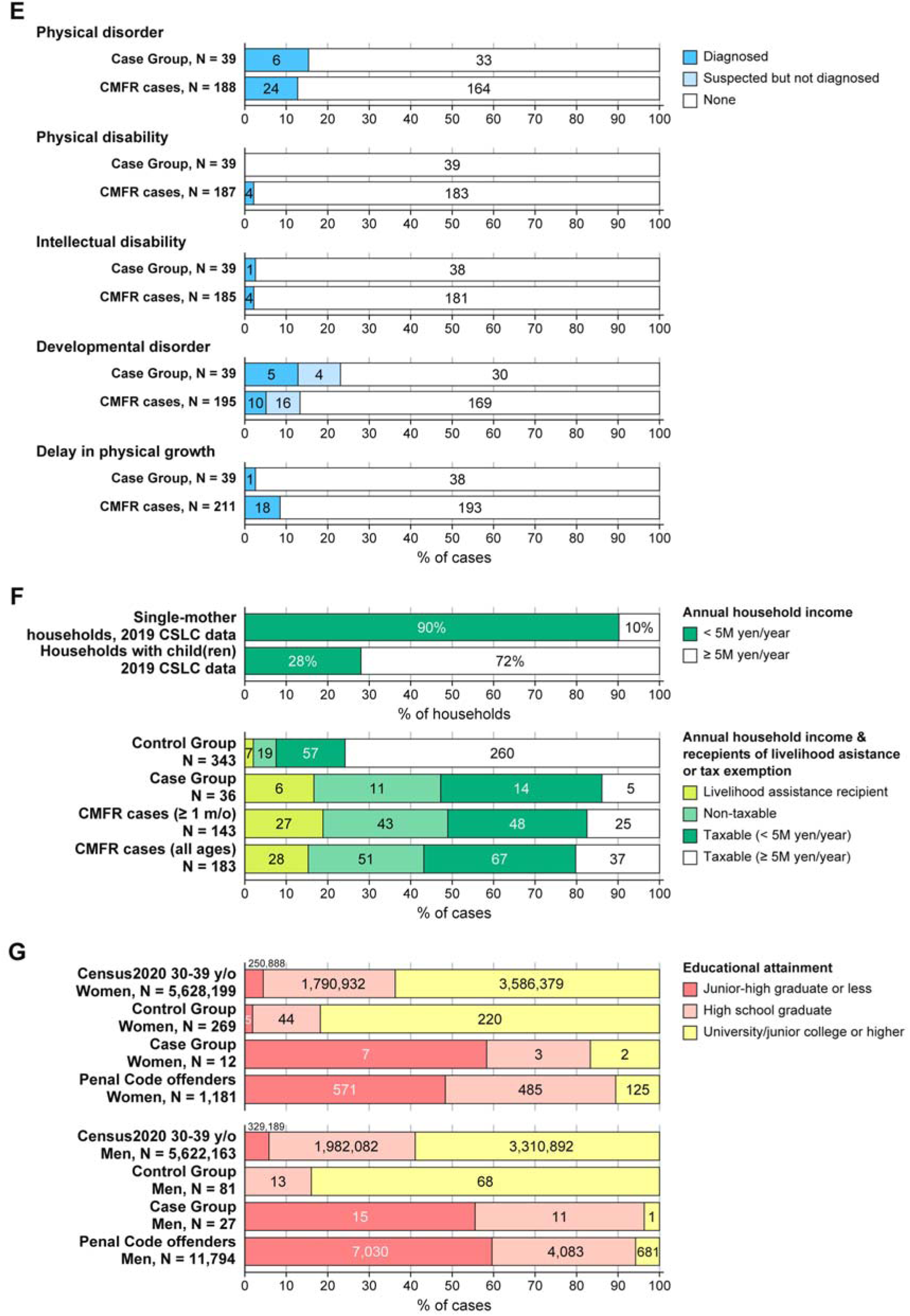
Comparisons of the participants to the relevant populations in the government statistics. **A-E**. The Case group is compared with cases in the Child Maltreatment Fatality Reviews (CMFR cases) ^17^, excluding neonaticide (≥ 1m/o, the cases with 0-month-old victims). The latter group is more similar to our Case group than the CMFR group including victims of all ages, because the rates of name disclosure and conviction are lower in the neonaticide cases, often performed only by the mother. Neonaticide accounts for 23% of the CMFR cases, of which 88.3% were performed by a young genetic mother alone. **A.** Types of abuse. Our Case group had a significantly lower proportion of neglect cases than the CMFR cases excluding neonaticide (P = .025, OR = 0.35 [0.13, 0.91]). **B.** Age distribution of victimized children. No significant differences between our Case group and the CMFR cases excluding neonaticide (mean: Student’s t-test, t = -0.41, df = 431, P = .685; distribution: two-sample Kolmogorov-Smirnov test, D = 0.14, P = .501). **C.** The relations between the victimized child and the principal perpetrators of the case. Compared to the CMFR cases excluding neonaticide, our Case group had fewer cases committed by a genetic mother alone (P < .001, OR = 0.18 [0.07, 0.45]) and more cases by a genetic mother with a non-kin man (stepfather or de facto husband; P < .001, OR = 6.30 [2.60, 15.10]). **D.** The types of caregivers with whom the target children lived at the time of the incident or the target period. Compared to the CMFR cases excluding neonaticide, fewer children in our Case group had lived with both of their genetic parents (P = .02, OR = 0.42 [0.21, 0.85]) and more lived with a non-kin caregiver (P < .00, OR = 7.13 [3.30, 15.45]). The difference in the proportion of single-parenting caregivers was significant but small (P = .049, OR = 0.30 [0.07, 0.94]). According to the 2020 Census, 21.5% and 4.1% of households with children in Japan were mother-child and father-child households, respectively ^21^. **E.** Health and developmental issues in the victimized children. No significant difference was found between our Case group and the CMFR cases across categories. **F.** Economic situation of the households. The top two bars represent the annual income of the single-mother households and all households with child(ren) in Japan from the 2019 Comprehensive Survey of Living Conditions (CSLC). None of these categories was significantly different between our Case group and the CMFR cases. Except for the CSLC data, households with annual income < 5 million yen were further categorized by whether they were receiving livelihood assistance under the Public Assistance Act or exemption from the Inhabitant Tax. **G.** Educational attainment of our participants, the Japanese Penal Code offenders who were newly imprisoned in 2016 ^22^, and the 2020 Census population. None of the categories was significantly different between our Case group and the Penal Code offenders, regardless of gender. In our Case group, 58.3% of women and 55.6% of men didn’t have a high-school diploma, and neither did 48.3% of women and 60.0% of men in the Penal Code offenders. These proportions are higher than the general Japanese population aged 30–39 years (Census 2020, women: 4.5%, men: 5.9%). Our Control group contained more participants with higher educational backgrounds than the Census 2020 population (university/junior college or higher; w: P < .000, OR = 2.56 [1.87, 3.53], m: P < .000, OR = 3.65 [2.01, 6.92]. Our control group appears to be biased toward higher education, possibly due to the research design requiring substantial reading and handwriting, as well as multiple communications via postal mail. Note that the Census 2020 population includes individuals with intellectual and other disabilities. Except for B, 2×2 Fisher’s exact test was used. M/o: months old. Due to differing categorizations, CMFR datasets varied across panels: A, B: 2008–2018 fiscal year (FY). C: 2010–2018 FY. D, F: 2012–2018 FY. E: 2013–2018 FY. The CSLC data analyzed 22,288 households in total for the Income Questionnaire Survey. The term “child” in this survey refers to an unmarried person aged < 18, and “mother-child(ren) household” refers to a household consisting of a woman aged <r 65 without a spouse and her child(ren) aged < 20, including adopted child(ren). Percentages of livelihood assistance and tax exemption recipients were not available in this dataset.

**Figure S2.**
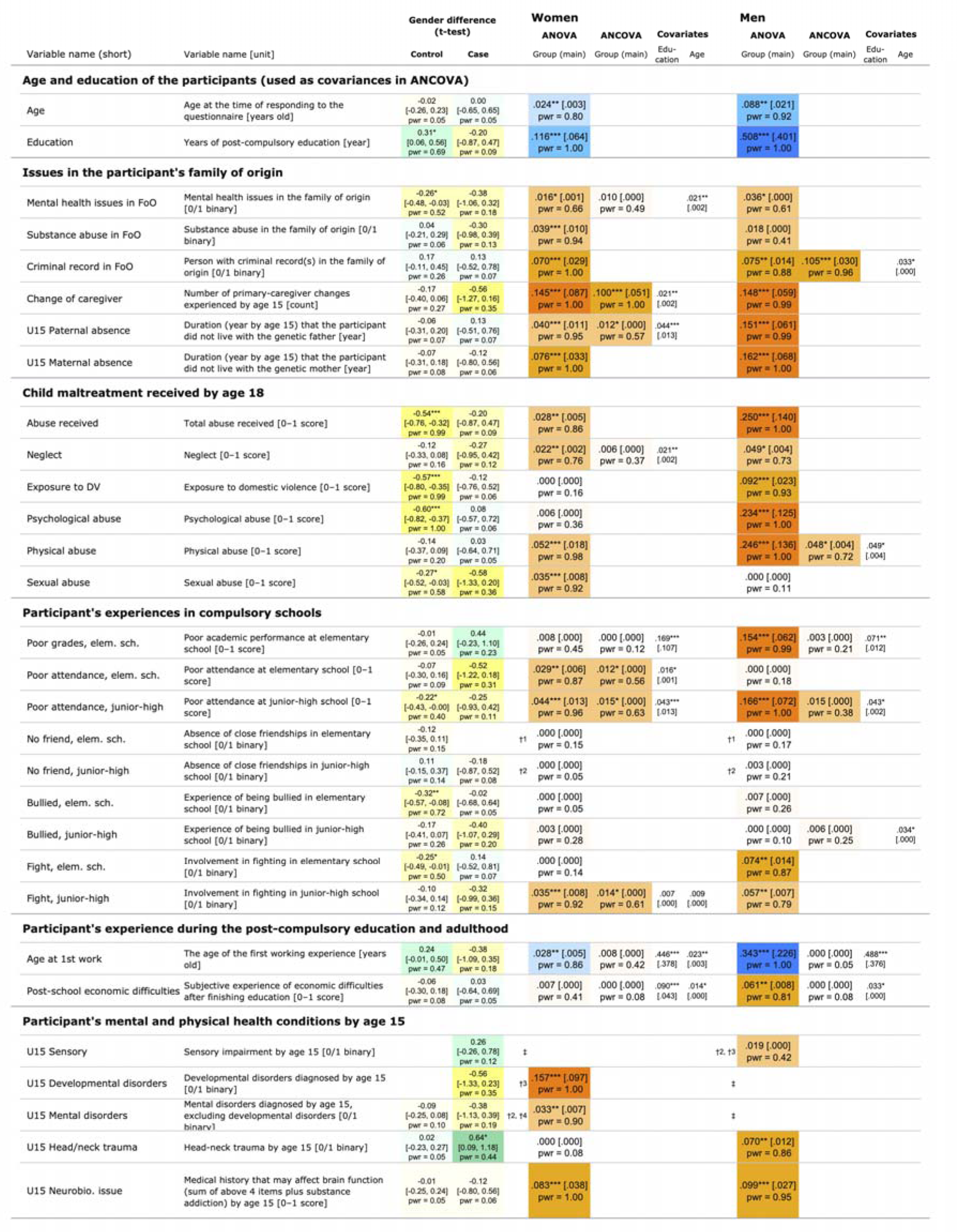
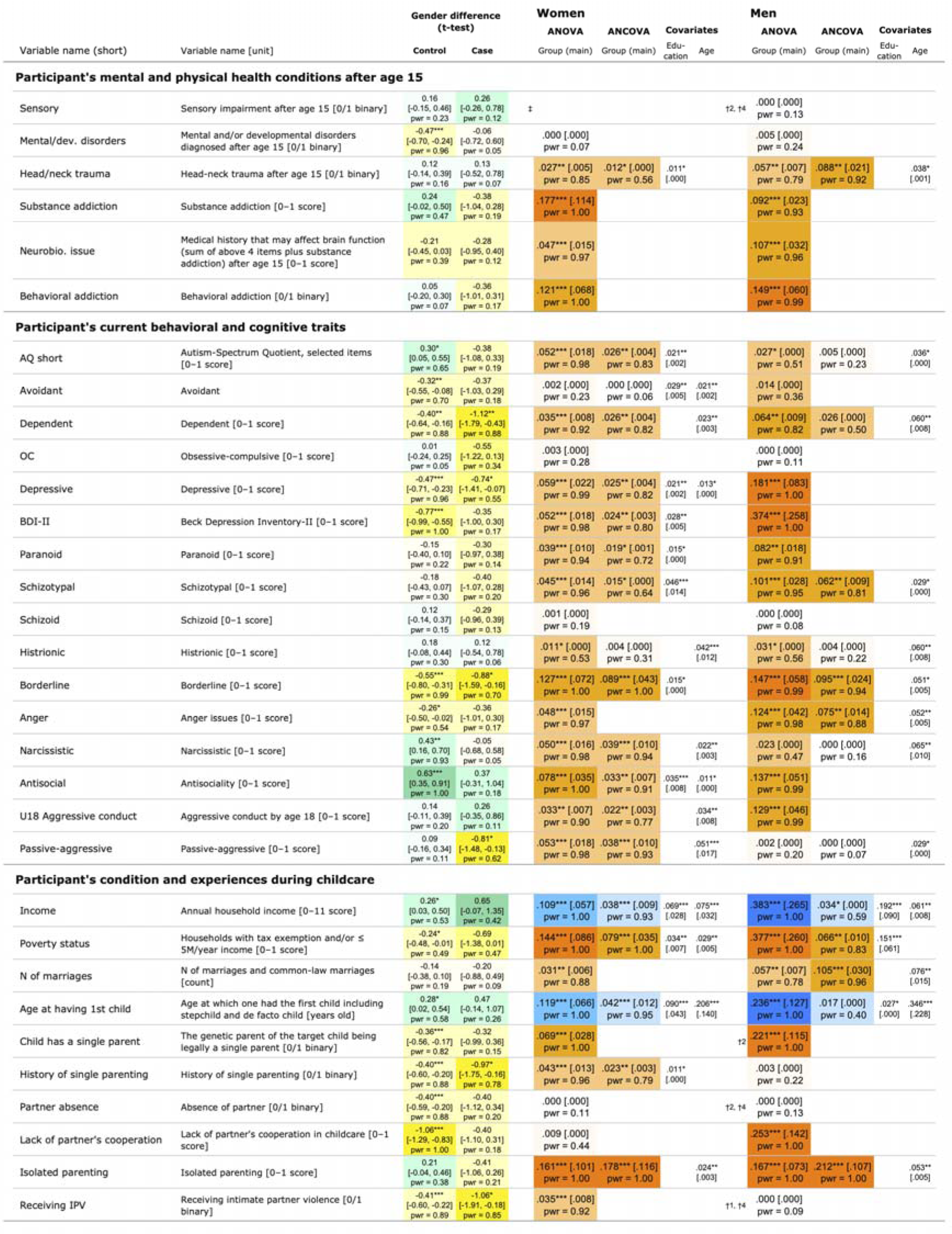
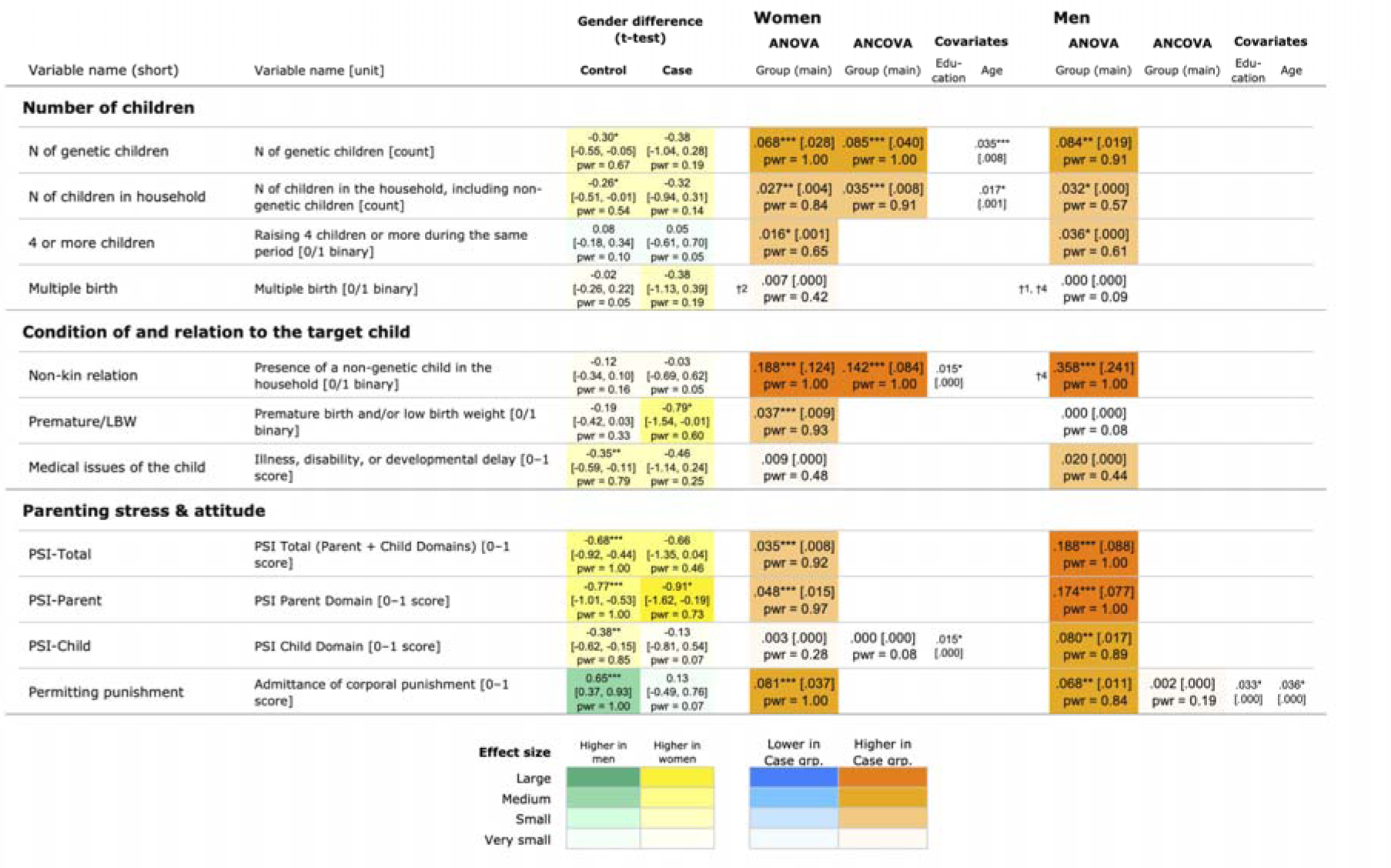
Effect sizes of gender and group comparisons on the background factors of child maltreatment. 70 independent variables and 3 variables generated as a composite of other variables were analyzed. Gender difference of 73 parameters within each group was analyzed using Welch’s t-test, and the corresponding effect sizes are presented as Hedges’ g. Group differences of 73 parameters within each gender were analyzed using ANOVA and ANCOVA, and effect sizes for the main effect (Group) and for the covariances (*age* and *education*) are presented as _ω_^2^ (omega-squared). Cell colors represent the interpretation of effect sizes, as indicated below the table. Values in the brackets denote 95% confidence intervals (two-sided for Hedges’ g and one-sided for _ω_^2^). Asterisks denote statistical significance for the main effect and covariates (* P < .05, ** P < .01, *** P < .001). For binary variables in which no or only one participant met the criterion in a group, daggers denote the following cases: †1, no applicable participant in the Case group. †2, only one applicable participant in the Case group. †3, no applicable participant in the Control group. †4, only one applicable participant in the Control group. ‡, no applicable participant in either group..

**Figure S3.**
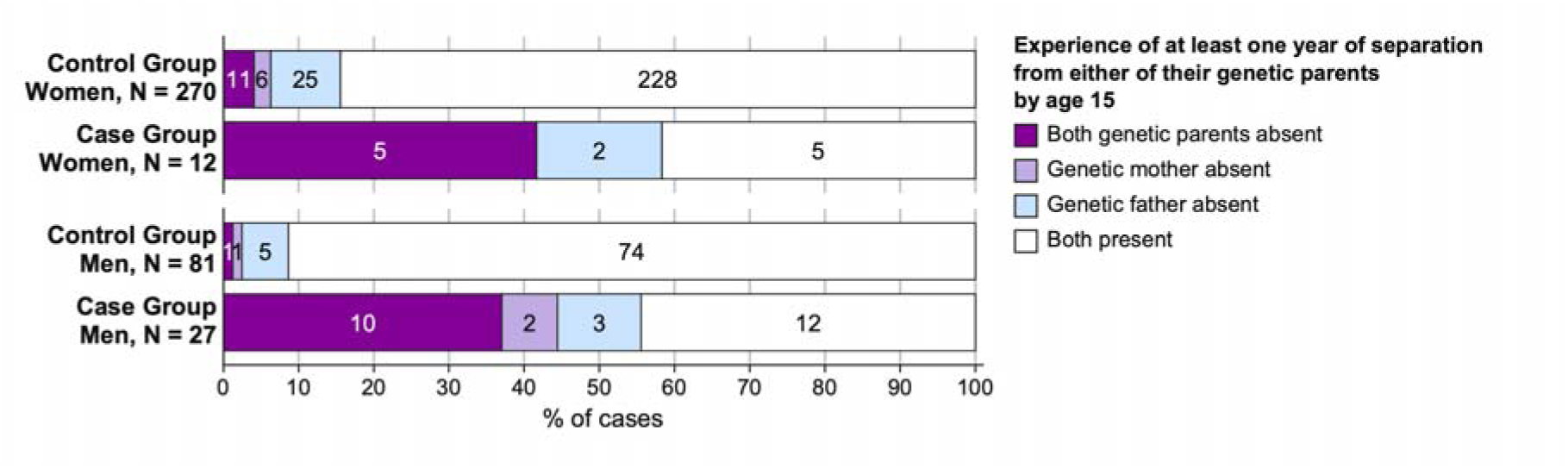
Absence of the genetic parents during childhood of participants. Absence of the participants’ genetic parents in their family of origin. The categories were determined by whether there was any year before age 16 in which both or either genetic parent was absent from the family. Compared to the Control group, the Case group had a significantly higher proportion of individuals who experienced the absence of both genetic parents (w P < .001, OR = 16.35 [4.41, 63.39]; m P < .001, OR 44.86 [6.64, 1017.8]). Conversely, the Case group had a significantly lower proportion of individuals whose genetic parents were both present throughout the 16 years of childhood (w P = .001, OR = 0.13 [0.04, 0.45]; m P < .001, OR 0.08 [0.03, 0.23]).

**Figure S4.**
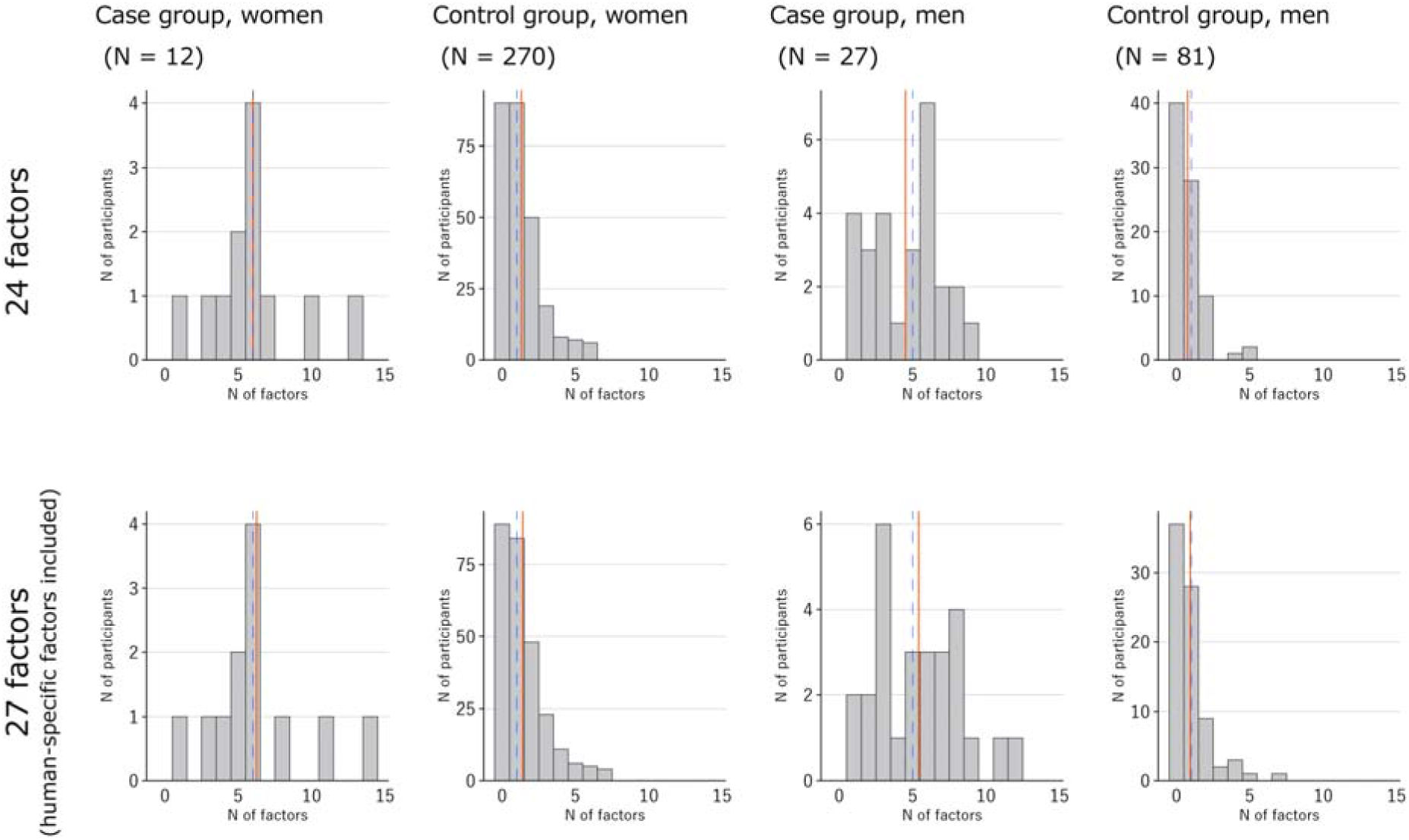
Distribution of the number of risk factors per participant. Analyses of the 24 key parameters, selected as human counterparts to established risk factors for mammalian parenting across five domains ^12^ (Table 1), showed that the Case participants accumulated more risk factors than Controls. When human-specific maltreatment types (*exposure to DV*, *psychological abuse*, and *sexual abuse*) were included, the disparities remained significant. In the histogram, red and blue lines denote the mean and the median, respectively.

**Figure S5.**
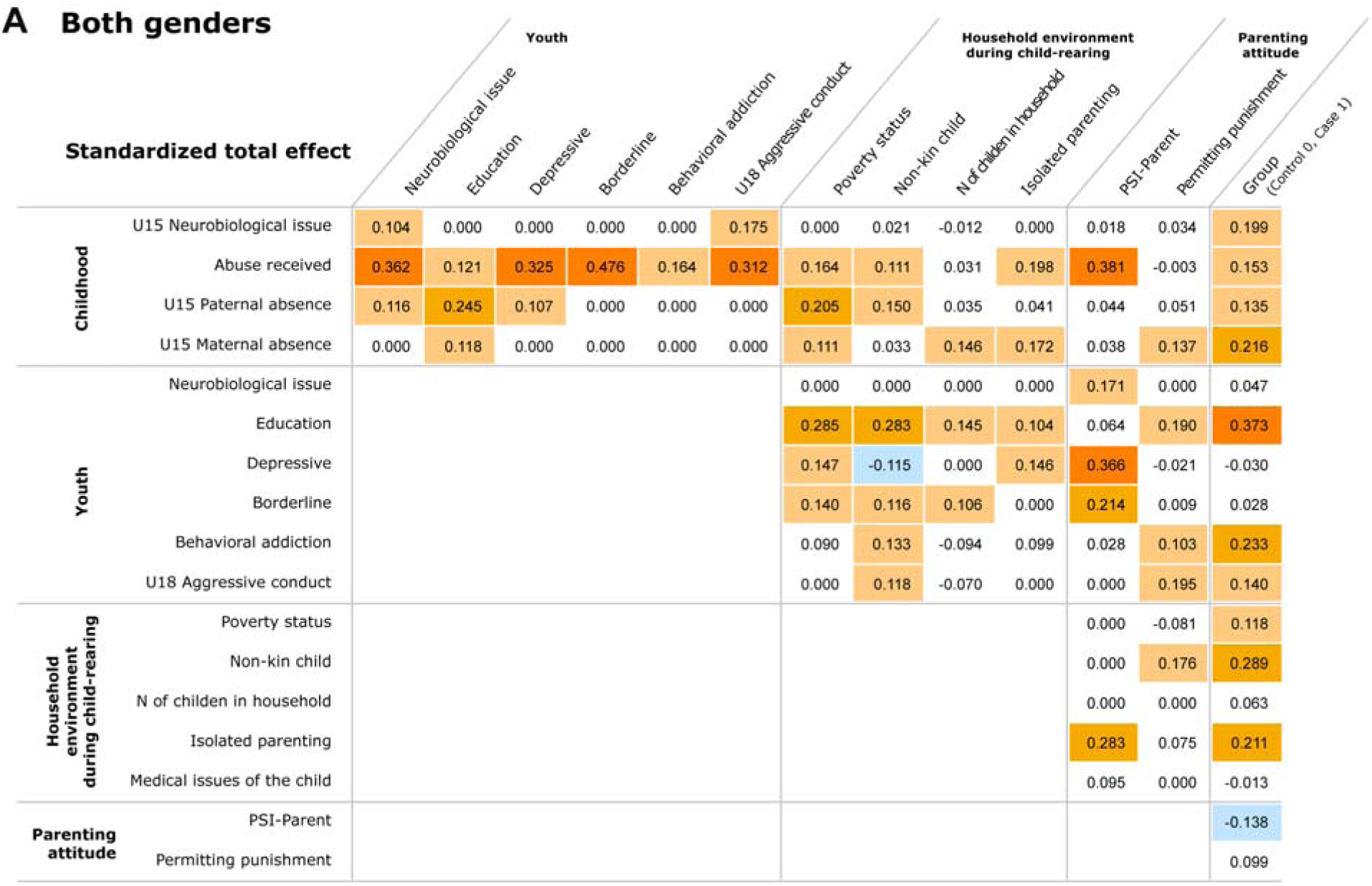
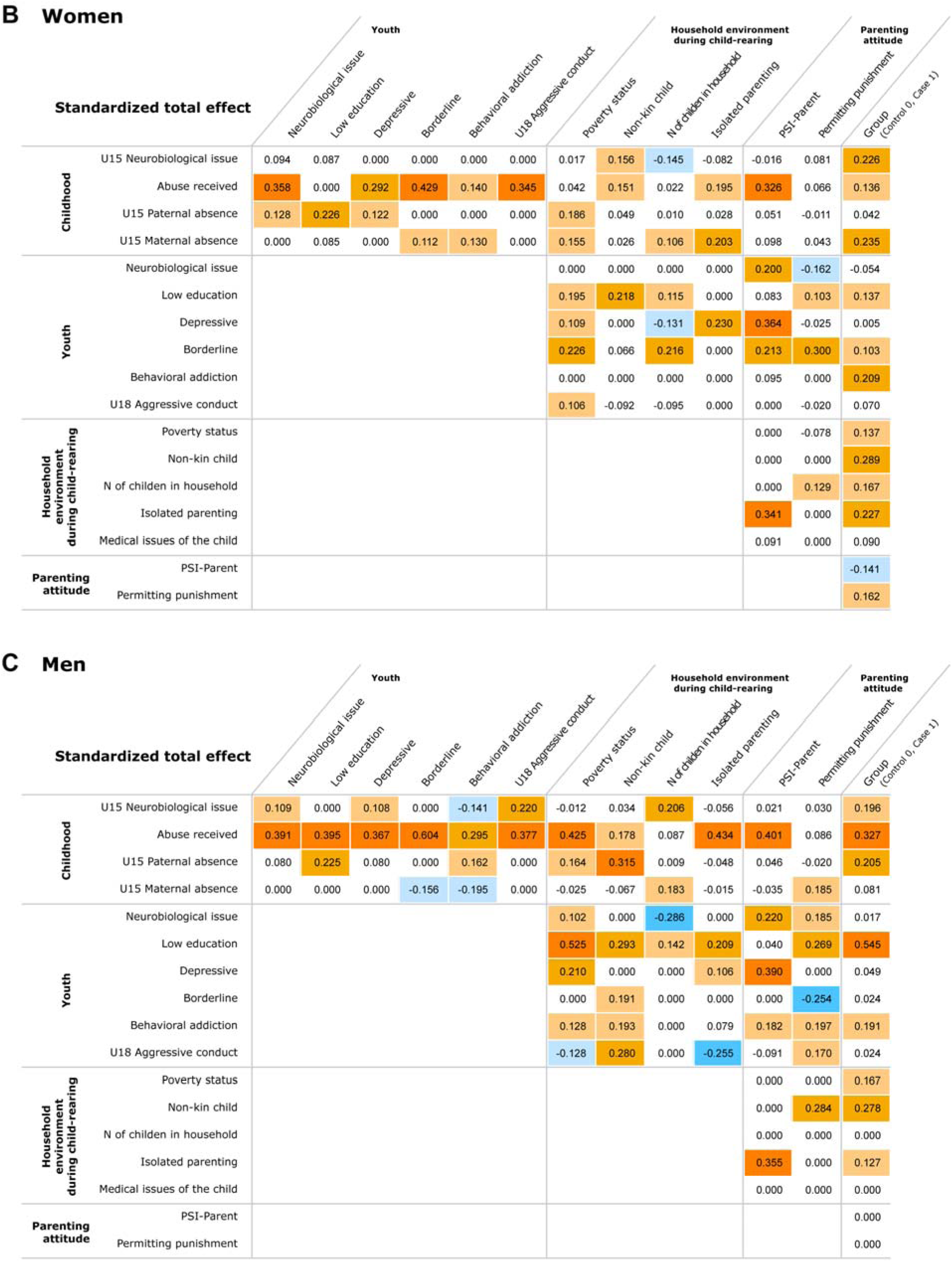
Standardized total effects of the final models in each gender. Standardized total effects of the final model using three separate datasets. **A**, both genders. **B**, women only. **C**, men only. Cell colors correspond to the legend in Fig. 5A.

